# Geographical and Gender Diversity in Cochrane and non-Cochrane Reviews Authorship: A Meta-Research Study

**DOI:** 10.1101/2024.03.23.24304672

**Authors:** Ahmad Sofi-Mahmudi, Jana Stojanova, Elpida Vounzoulaki, Eve Tomlinson, Ana Beatriz-Pizarro, Sahar Khademioore, Etienne Ngeh, Amin Sharifan, Lucy Elauteri Mrema, Alexis Ceecee Britten-Jones, Santiago Castiello-de Obeso, Vivian A. Welch, Lawrence Mbuagbaw, Peter Tugwell

**Author notes:** **Corresponding author:** Ahmad Sofi-Mahmudi; **Address**: MDCL 2109, McMaster University, 1280 Main Street West, Hamilton, ON L8S 4K1, Canada; **Email:**. **Conflict of interest disclosure:** AS-M, EV, AB-P, EN, AS, LEM, and SC-dO are members of the steering group of the Early Career Professionals Network in Cochrane. ET, VAW, LM, and PT are members of the Health Equity Thematic Group in Cochrane. AB-P and PT are members of the Editorial Board of Cochrane. VAW declares funding from CIHR-PHAC Applied Public Health Chair. **Funding disclosure:** This study did not receive any funding.

## Abstract

**Background:** Cochrane is a recognized source of quality evidence that informs health-related decisions. As an organization, it represents a global network of diverse stakeholders. Cochrane’s key organizational values include diversity and inclusion, to enable wide participation and promote access. However, the diversity of Cochrane review authorship has not been well summarized.

**Objective:** The aim of this study was to examine the distribution of country, region, language, and gender diversity in the authorship of Cochrane and non-Cochrane systematic reviews.

**Methods:** We retrieved all published articles from the Cochrane Library (until November 6, 2023)—a web crawling technique that extracted pre-specified data fields, including publication date, review type, and author affiliations. We used E-utility calls to capture the data for non-Cochrane systematic reviews. We determined the country and region of affiliations and the gender of the first, corresponding, and last authors for Cochrane reviews, as well as the country and region of affiliations and the gender of the first authors for non-Cochrane reviews. Trends in geographical and gender diversity over time were evaluated using logistic regression. Fisher’s exact test was used for comparisons. The diversity of first authors between Cochrane and non-Cochrane reviews was explored through visual presentation, Pearson’s product-moment correlation, and the Granger Causality Test. We used R for data collection and analysis.

**Results:** A total of 22681 citations were retrieved. The United Kingdom had the highest first-author representation (33.2%), followed by Australia (11.6%) and the United States (7.0%). We observed an increase in the proportion of first authors from non-English speaking countries, from 16.7% in 1996 to 42.8% in 2023. Female first authorship increased steadily, from 15.0% in 1996 to 55.6% in 2023. The proportion of first authors from lower-and-middle-income countries (LMICs) was highest in 2012 at 23.2%. Since then, it has decreased to 18.4% in 2023. Similarly, the proportion of last authors from LMICs decreased over time (25.0% in 1996 vs. 16.2% in 2023). Among review groups, Sexually Transmitted Infections and Consumers and Communication were the most and least diverse groups with 68.1% and 1.6% of first authors from LMICs, respectively. In terms of gender diversity, Fertility Regulation had the highest percentage of female first authors (72.1%). Urology (28.1%) had the lowest percentage of female first authors. In 2023, over half of the non-Cochrane reviews had first authors from non-English-speaking countries (n=14,589, 56.9%), 50.8% (n=13,014) had first authors from LMICs, and 42.3% (n=10,841) had female first authors. The Pearson’s product-moment correlations between Cochrane and non-Cochrane reviews’ trends were 0.265 (P=0.450) for LMICs, 0.823 (P<0.001) for non-English speaking, 0.634 (P<0.001) Spanish-speaking, and 0.829 (P<0.001) for female first authorship.

**Conclusion:** Overall, this study found positive trends, with an increase in first authorship by individuals who were female and from non-English speaking countries. However, the representation of first authors from LMICs decreased. Future research could further explore these trends, identifying potential barriers influencing access and participation of individuals and groups and assessing strategies that help promote diversity and inclusion.

## Introduction

Global health challenges transcend geographical boundaries. For over 30 years, Cochrane has brought together diverse researchers and stakeholders within a large global network, with the aim of producing high-quality systematic reviews that address important challenges in healthcare (1). These comprehensive reviews are pivotal in guiding clinical practice, policy development, and research agendas (2). A further organizational aim is the translation of research findings, and Cochrane supports global reach in research translation through a network of Geographic Groups, currently representing 54 countries (3).

With origins in the United Kingdom, and early membership predominantly representing anglophone countries, Cochrane has grown substantially as an organization with wide global reach. Of the 137 Cochrane Geographic Groups, 114 are from countries where English is not the primary spoken language (3). Cochrane’s vision is “a world of better health for all people where decisions about health and care are informed by high quality evidence” with the aim to make evidence accessible to all (1). Collaboration is one of the core values in the organizational strategy (4) and the organization has multiple avenues for participation and inclusion, including membership (predominantly reflecting status as a recent author) and supporter (which may involve active participation through initiatives such as crowd-sourced screening, among many other initiatives) (5). Membership in leadership structures, such as the Governing Board and Council, is attained through member vote, and these entities are structured to support wide representation. Despite this, collated feedback from over 1300 people over the world in a “diversity and inclusion listening and learning exercise,” reported by Cochrane in 2022, highlighted that Cochrane is not as diverse and inclusive as it could be, and has work to do to address systemic institutional biases in its systems, processes, and attitudes (6).

Cochrane’s key output is the systematic review. Agendas regarding strategic topics for reviews are set by Cochrane Review Groups (CRGs), a majority of which are based in high-income, anglophone countries (6). While some CRGs undertake priority-setting processes involving stakeholders to determine research priorities (7), in line with the 2019 Cochrane Priority Setting Guidance (7), it is unclear how common this is, and Cochrane leadership has recognized that this process often does not have a global focus (8). Furthermore, research has found that Cochrane Reviews tend to be authored primarily by individuals from high-income countries, with limited representation from low- and middle-income countries (9–12). There is also a gender imbalance, with women underrepresented among Cochrane Review authors (13,14). This is an issue, as broad representation of authors from different countries, regions, languages, and genders brings a wider range of experiences, knowledge, and perspectives to the review process, enriching the synthesis and interpretation of evidence (15). This diversity is likely to help to ensure that Cochrane Reviews consider issues of health equity and in turn that the findings apply to a wide range of populations and healthcare settings, or that they specifically address the unique needs and circumstances of disadvantaged groups (16).

Publications on this topic today have focused on narrow topics such as hematology (9), eyes and vision (10), gastroenterology (11), cardiology (13), and general surgery (14), or a specific geographical location (e.g., sub-Saharan Africa (17)). Thus, this meta-research study aims to assess the distribution of country, region, language, and gender diversity in Cochrane and non-Cochrane reviews’ authorship. We compared income status (high vs lower-and-middle-income countries (LMICs)), English-speaking countries (vs others), and the gender of the first, last, and corresponding authors. Given that approximately a third of Cochrane Geographic groups are based in Spanish-speaking countries, and have a dedicated conglomerate, Cochrane Iberoamerica, we also evaluated representation from these countries. Further, we investigated diversity in the first authors of non-Cochrane reviews and compared results with those from Cochrane reviews. A fully automated and reproducible approach was applied to systematically extract and analyze author information from both sets of reviews (Cochrane and non-Cochrane).

## Methods

The study protocol was published on the Open Science Framework (OSF) website (https://osf.io/bxj2e). Deviations from the protocol are detailed in Appendix 1. All datasets and codes of workflows used in this study are publically available (OSF: https://osf.io/fv5ys, GitHub: https://github.com/choxos/cochraneauthors). To ensure transparency and facilitate the reproducibility of our analyses, a PDF document containing the codes and corresponding outputs is provided in Appendix 2.

### Data sources and retrieval

All reviews published by Cochrane on the Cochrane Library website (cochranelibrary.com) were retrieved (up to November 6, 2023). Since the Cochrane Library provides only the latest version of reviews in their search interface, links to all review versions were automatically created using standard patterns for the digital object identifier (DOI). A typical DOI has the format: “10.1002/14651858.CD” + Review ID + “.pubN”.

The first version is usually the protocol, and does not include “.pubN”. The N represents subsequent protocol versions, e.g. “.pub2” for version 2. All the possible DOIs were created automatically and used for the final extract of Review titles.

We applied a web crawling technique to extract pre-specified data fields for each review from their dedicated information page on the Cochrane Library website (including date, review type, review stage, review group, author position, and author affiliation for all authors). The URL of the review information page was structured as follows: “https://www.cochranelibrary.com/cdsr/doi/” + DOI + “/information”

Authorship position/role was determined (first, last, corresponding). Affiliations were categorized according to country and World Bank economic status. We then categorized the country of the first, corresponding, and last authors in three different ways: (A) high-income vs. LMICs, (B) high-income English-speaking vs. non-English-speaking, and (C) Spanish-speaking vs. non-Spanish-speaking. The list of the countries in each of these categories is available in Appendix 3.

The authors’ gender was attributed using the World Gender Name Dictionary 2.0 (18). This database includes approximately 3.5 million names from different languages across the world, and the probability that a name is considered male or female is higher. For this study, we considered the higher probability as the definitive gender and assigned a dichotomous gender variable for all authors.

Review updates can have the same author composition as the previous version, although there are deviations (for example, see (19–23) and their previous versions). Also, in this study, the unit of analysis is a published paper and not a project. Therefore, we included all the updates of a review in our analyses.

All non-Cochrane systematic reviews were retrieved from PubMed using the following search query:(“Systematic Review”[PT]) NOT (“The Cochrane database of systematic reviews”[Journal], using E-utility calls (24) from 1996 to 2023 (to be comparable with Cochrane reviews). We extracted the PMID, publication date, name, and affiliation of the first author for each review and applied the approach detailed above to ascertain the gender, country, and region.

### Analysis

We used R (25) for data extraction, data processing, analysis, and reporting. Searching and data gathering for non-Cochrane review was done using a bash script available in Appendix 2. Web scraping was done using the *rvest* package (26). Trends over time were reported using descriptive tabulations and graphical illustrations using the *ggplot2* package (27). We used logistic regression to explore whether geographical and gender diversity has changed over the years. We also used a random intercept generalized linear model to investigate the trend of geographical and gender diversity among different review groups. To compare the geographical and gender differences between the first, corresponding, and last author between review groups, we performed Fisher’s exact test with 2000 replicates.

To compare the trend of first authorship diversity between Cochrane and non-Cochrane reviews, alongside visual presentation and Pearson’s product-moment correlation, we also used the Granger Causality Test (28,29). This test assesses whether past values of one time series can predict future values of another. The null hypothesis is that one time series does not cause the other.

### Validation

To validate the accuracy of the automatically extracted data, we randomly sampled 5% of the dataset and manually verified the names and countries of the first, corresponding, and last authors. This sample is available in Appendix 4.

## Results

### Overall perspective

We extracted 22,681 articles, of which 9,153 (40.4%) were the most recent review version and 7,157 (31.6%) were protocols. The annual number of published articles (from 1995 to November 2023) is presented in Appendix 2 (mean=782.1, standard deviation=446.02). Publications peaked in 2012 (n=1,508) and the most recent total was 376 (in 2023). Most articles represented interventional reviews (n=21,965, 96.8%). Diagnostic reviews (n=358, 1.6%) and overviews (n=140, 0.6%) had minor representation.

The Cochrane Review Groups with the highest number of published reviews were Pregnancy and Childbirth (now closed) (n=1,634, 7.2%), Neonatal (n=1,118, 4.9%), and Airways (n=873, 3.8%). Lower representation was apparent for Sexually Transmitted Infections (n=47, 0.2%), Methodology (n=104, 0.5%), and Work (n=108, 0.5%) Groups. Twenty-three reviews were collaborations between two review groups. The yearly trend of the number of reviews by each group is available in Appendix 5.

### Geographical diversity

Similar trends were observed across different author types (Table 1). First authors were from a greater number of countries compared to the last authors (102 vs. 93). Across author types, 107 countries were represented. Regardless of the author type, most authors were from high-income and English-speaking countries (approximately 80% and 60%, respectively). Most authors were from the United Kingdom, Australia, and the United States (approximately one-third, 10%, and 7% respectively). A World heat map of the countries based on the number of authors is presented in Figure 1. The detailed information about the countries is available in Appendix 6.

**Figure 1.**
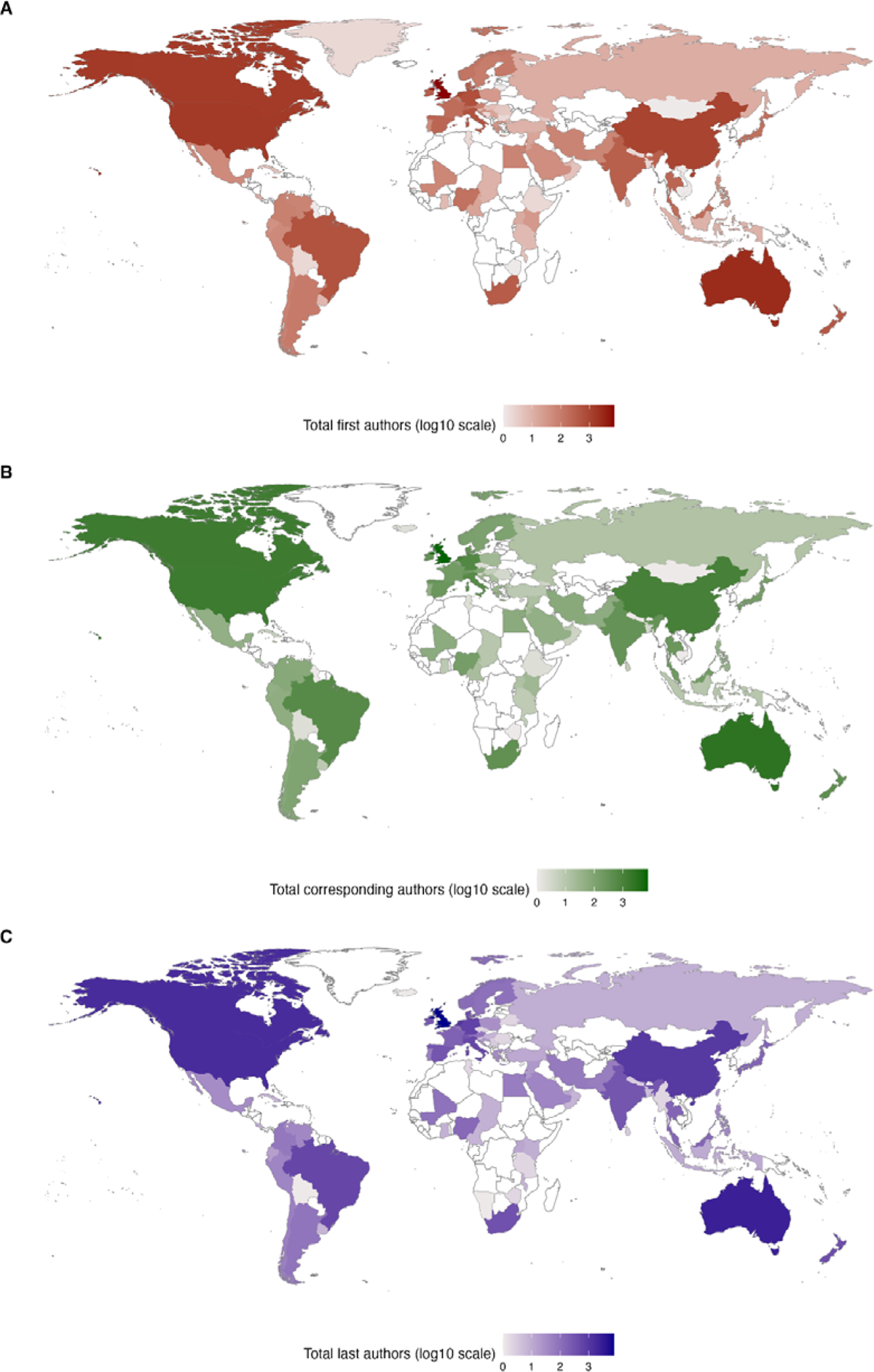
World heat map of the countries based on the number of (A) first, (B) corresponding, and (C) last authors (in log10 scale).

**Table 1.** Summary of the number of Cochrane authors by each diversity index (from 22,681 reviews).

|  | **First authors** | **Corresponding authors** | **Last authors** |
| --- | --- | --- | --- |
| Number of countries | 102 | 98 | 94 |
| Number of affiliations where a country could not be identified | 318 (1.4%) | 112 (0.5%) | 442 (1.9%) |
| Number of countries represented by a sole review | 12 (11.8%) | 12 (12.2%) | 8 (8.5%) |
| **Most represented countries** |  |  |  |
| United Kingdom | 7,426 (33.2%) | 7,412 (33.2%) | 7,720 (34.7%) |
| Australia | 2,595 (11.6%) | 2,619 (11.7%) | 2,640 (11.9%) |
| United States | 1,559 (7.0%) | 1,537 (6.9%) | 1,626 (7.3%) |
| **Income status** |  |  |  |
| High-income | 18,195 (81.9%) | 18,234 (80.4%) | 18,682 (82.4%) |
| Lower-and-middle-income | 4,016 (18.1%) | 4,446 (19.06%) | 3,998 (17.6%) |
| **English speaking country** |  |  |  |
| Yes | 13,866 (62.0%) | 13,871 (62.1%) | 14,252 (64.1%) |
| No | 8,497 (38.0%) | 8,469 (37.9%) | 7,987 (35.9%) |
| **Spanish speaking country** |  |  |  |
| Yes | 753 (3.4%) | 743 (3.3%) | 651 (2.9%) |
| No | 21,610 (96.6%) | 21,597 (96.7%) | 21,588 (97.1) |
| **Gender (by first name)** |  |  |  |
| Female | 10,545 (50.8%) | 9,947 (48.1%) | 7,804 (37.3%) |
| Male | 10,207 (49.2) | 10,720 (51.9%) | 13,106 (62.7%) |

Author representation by income status (high/LMIC) and language (English/non-English speaking and Spanish/non-Spanish speaking) over time and by author type are presented in Figure 2. In a given year, LMIC representation was at most 26.7% (first authors in 1996), and non-English country representation was at most 43.8% (corresponding authors in 2020). The first authors exhibited a greater representation of LMIC and non-English countries than the last authors. Following initial growth, the rate plateaued from about 2009 for non-English representation, and exhibited a decrease after 2012 for LMIC status. The results of the logistic regression modelling showed that the effect of year on the proportion of articles in each diversity index varied between 0.997 and 1.029 with *P*-values <0.001 except for Spanish-speaking and the last author from LMICs models (Appendix 7).

**Figure 2.**
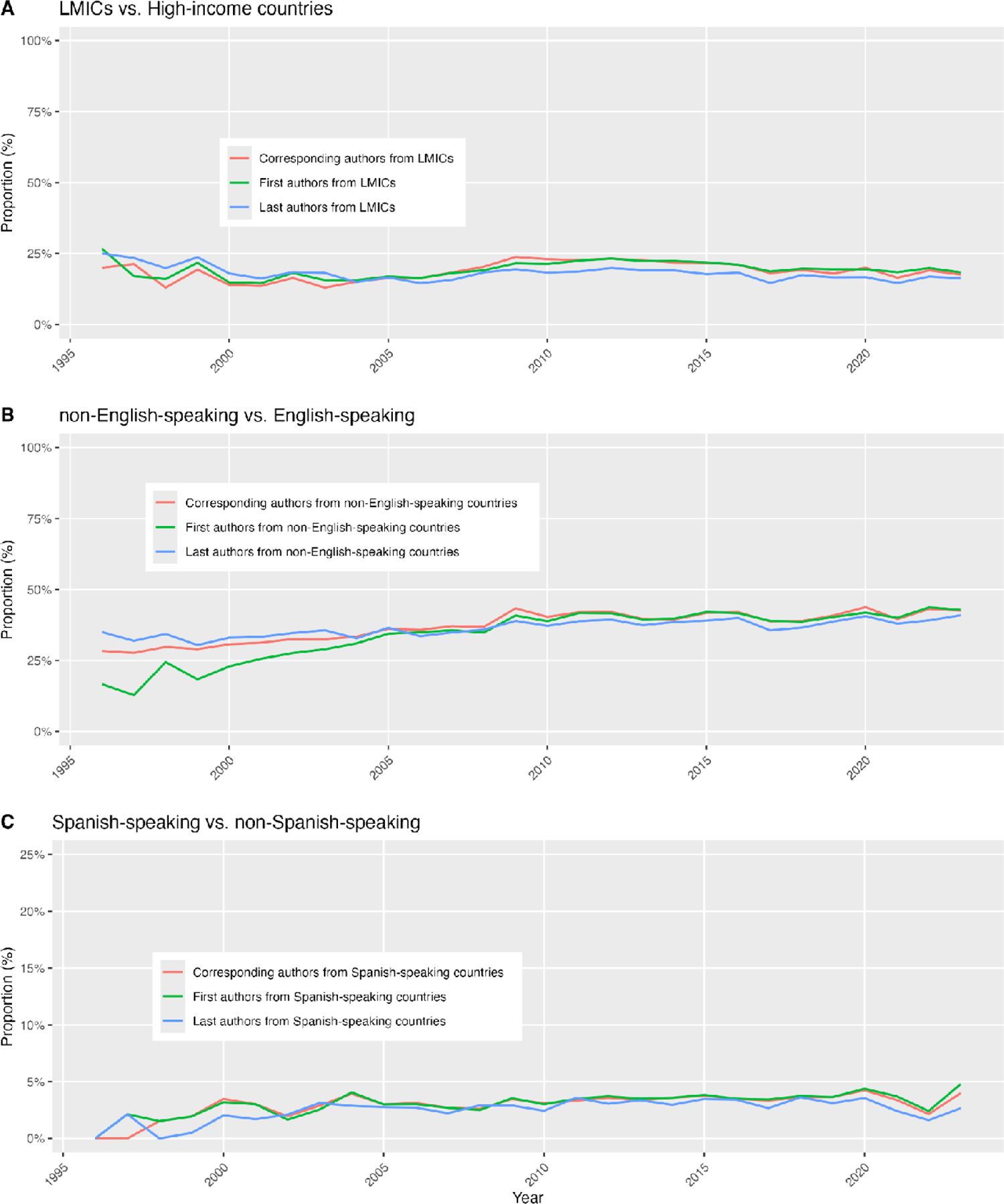
Region diversity in Cochrane Reviews authorship. (A) low-and-middle-income countries (LMICs) vs. non-LMICs. (B) non-English-speaking vs. English-speaking. (C) non-Spanish-speaking vs. Spanish-speaking. Please note that the y-axis for C is magnified to 0%-25%.

### Gender diversity

Over time, female first authorship increased from about a quarter in 1997 (27.7%) to more than half in 2023 (50.8%). Percentages of female corresponding and last authors likewise increased; however, growth was less pronounced for the last authors (39.4% in 2023; Figure 3). Logistic regression modelling showed that the coefficient for the year ranged between 1.015 and 1.030 with *P*-values <0.001 (Appendix 7).

**Figure 3.**
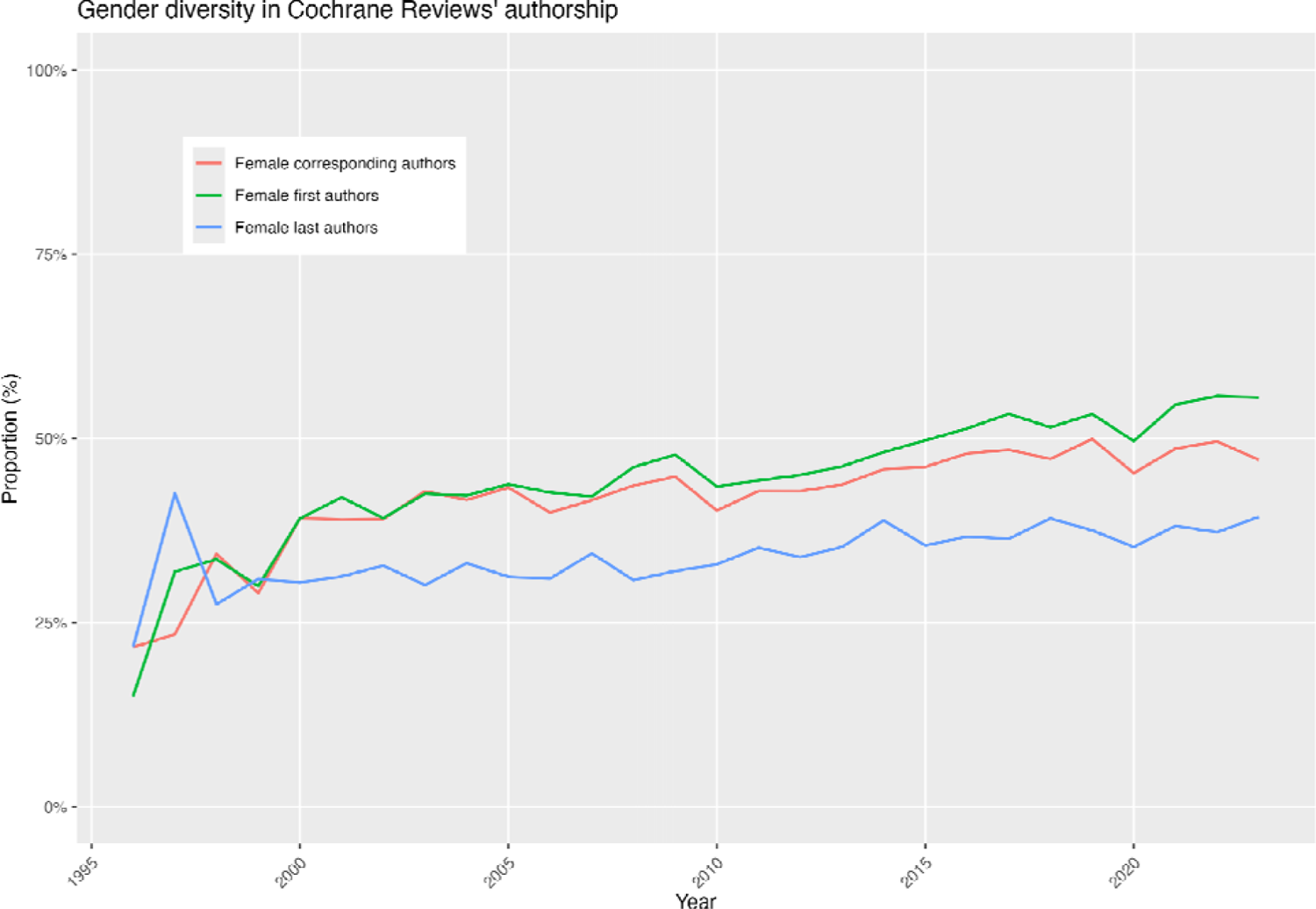
Gender diversity in Cochrane Reviews authorship.

### Diversity among Review Groups

Across author categories, most CRGs had 10-25% reviews with authors from LMICs and 25-50% of reviews with authors from non-English speaking countries, across author categories (Table 2). Seven CRGs had less than 10% of reviews with first authors from LMICs, and seven CRGs had 10-20% of first authors from non-English countries. The Sexually Transmitted Infections Review Group had the highest proportion of reviews with authors from LMICs (first=68.1%, corresponding=66.0%, and last authors=57.4%). This was the only CRG with representation above 50% of LMICs for all three author categories. HIV/AIDS and Infectious Diseases had the next highest LMIC representation, with 50.6% and 41.1% of first authors from these countries. Childhood Cancer (n=118 reviews; now closed) had the highest proportion of reviews with authors from non-English-speaking countries (87% of the three author categories combined). Lower geographical diversity was observed in the Consumers and Communication group, with 1.6% and 13.3% of reviews with first authors from LMICs and non-English countries, respectively.

**Table 2.** Number of CRGs with indicated proportions of reviews with first, corresponding, and last authors, that are from LMICs, non-English speaking countries and Female. Proportion categories were selected to best represent the data.

| Proportion of reviews | First authors | Corresponding authors | Last authors |
| --- | --- | --- | --- |
| LMICs |  |  |  |
| [0–10%) | 7 | 6 | 9 |
| [10–25%) | 33 | 33 | 35 |
| [25–50%) | 11 | 13 | 8 |
| [50–100%] | 2 | 1 | 1 |
| **Non-English-speaking countries** |  |  |  |
| [0–10%) | 0 | 0 | 0 |
| [10–25%) | 7 | 7 | 7 |
| [25–50%) | 30 | 30 | 33 |
| [50–100%] | 16 | 16 | 13 |
| **Female (by first name)** |  |  |  |
| [0–10%) | 0 | 0 | 0 |
| [10–25%) | 0 | 0 | 8 |
| [25–50%) | 20 | 26 | 38 |
| [50–100%] | 33 | 27 | 7 |

LMICs: Low-and-middle-income countries

Most CRGs had more than 50% of reviews with female first and corresponding authors, however, a majority of CRGs had 20-50% of reviews with female last authors, suggesting a ‘ceiling’ for this author category. Fertility Regulation had the highest percentage of female first authors (72.1%), followed by Consumers and Communication (69.1%), and Skin (66.6%). Lower representation of female first authors was observed for reviews from the Urology, Hepato-Biliary, and Colorectal Groups (less than 35%). Appendix 8 shows bar plots of geographical and gender diversity for each CRG and Appendix 9 presents percentages.

The results of Fisher’s exact test showed a *P*-value<0.001 for all the comparisons between review groups in terms of geographical and gender diversity. The random intercept generalized linear mixed-effects logistic models showed higher random effects variance for the effect of CRGs for Spanish-speaking models (about 1) and lower for female models (less than 0.2). Full details are available in Appendix 2 and 10.

### Comparison with non-Cochrane reviews

We retrieved 224,484 non-Cochrane systematic reviews, representing the period 1987 to 2024. Lack of information in first author affiliations precluded assigning country and language status for 41,266 (18.4%), and income region for 43,315 (19.3%) of reviews. Gender could not be assigned for 39,619 (17.6%) first authors.

In 1996, of 60 non-Cochrane reviews, none included first authors that were female, from LMICs, or from Spanish-speaking countries; however, 11 (18.3%) reviews had first authors from non-English-speaking countries. In 2023, of 25,629 reviews, over half had first authors from non-English-speaking countries (n=14,589, 56.9%), 50.8% (n=13,014) had first authors from LMICs and 42.3% (n=10,841) had female first authors (Figure 4).

**Figure 4.**
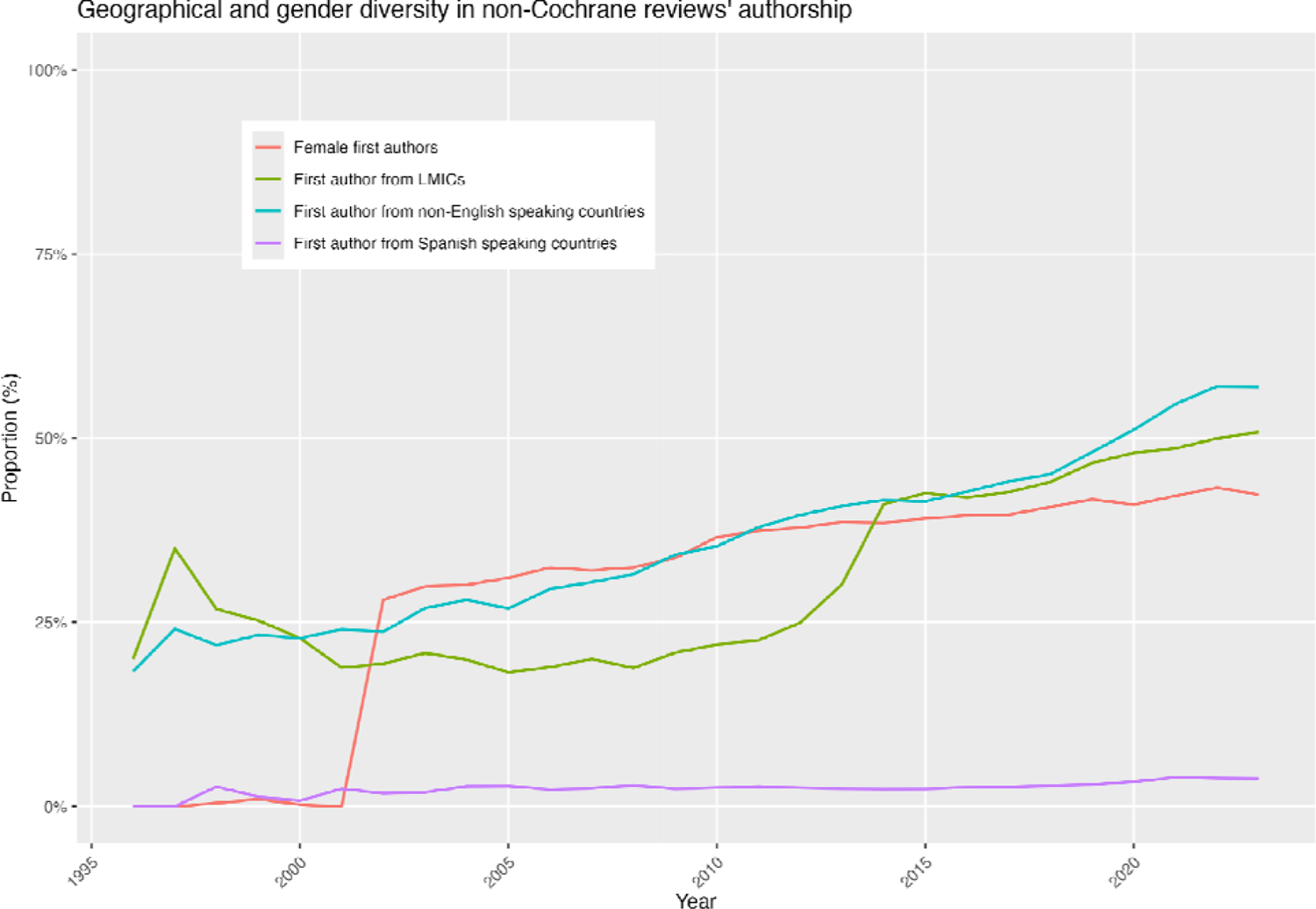
Geographical and gender diversity in the first authorship of non-Cochrane reviews

Trends over time exhibit notable differences compared to those observed with Cochrane reviews. One, the proportion of first authors in non-Cochrane reviews from LMICs is above 15% from 1996 to 2012, then exhibits a sharp increase to 41.0% in 2014, followed by a steady increase to 50.8% in 2023 (Figure 4); all in all, proportions over the period are higher than for Cochrane reviews, which are exhibit a relatively plateaued rate of growth (Figure 2). Two, the proportion of first authors of non-Cochrane reviews from non-English speaking countries starts at about 20% in 1996 and increases steadily to reach approximately 60% in 2023; the rate of growth for non-English speaking authors, regardless of author type, is lower in Cochrane reviews, where proportion does not reach 50% (Figure 2). Three, first authors from Spanish-speaking countries comprise less than 3% until the 2020s in which we see an increase to above 3% with 3.9% at its highest in 2021. Four, a sizable representation of female authorship in non-Cochrane reviews appears in 2002, starting at 28.0% (Figure 4), and continues to grow to 42.3% in 2023. In contrast, female representation in Cochrane reviews has been stable across author types since about the year 2000, and for first authors, it peaks in 2023 at 55.6% (Figure 2).

The results of the Granger Causality Test showed *P*-values of 0.062, 0.701, 0.483, and 0.499 when comparing LMIC, non-English speaking, Spanish-speaking, and female first authorship trends between Cochrane and non-Cochrane reviews. This means the non-Cochrane and Cochrane trends may not be predictive of each other. The Pearson’s product-moment correlations were 0.265 (*P*=0.450), 0.823 (*P*<0.001), 0.634 (*P*<0.001), and 0.829 (*P*<0.001), respectively.

### Validation

The validation sample contained 1134 reviews. There were no discrepancies between the automatic algorithm and manual checking (Appendix 4).

## Discussion

This work represents a comprehensive evaluation of geographic and gender diversity among Cochrane reviews since the first Cochrane review was published in 1995. We showed that the first author representation from LMICs peaked at 26.7% in 1996 and at 43.8% in 2020 for non-English speaking countries. Both categories exhibited growing rates through to approximately 2010, followed by plateau periods. From 2015, representation from LMICs exhibited a decline, decreasing to 16.2% in 2023. Overall, authors were predominantly from high-income and English-speaking countries, approximately 80% and 60%, respectively, and of English-speaking countries, predominantly from the UK (approximately one-third of all authors). Despite a very active community of researchers from Spanish-speaking countries in Cochrane, evidenced by a sizable proportion of Geographic Groups and a dedicated conglomerate (Cochrane Iberoamerica), author representation from Spanish-speaking countries was low. Non-Cochrane reviews were more diverse in having a higher proportion of first authors from LMICs (50.8% vs. 18.4% in 2023) and non-English-speaking countries (56.9% vs. 42.8% in 2023).

These results echo findings from previous work in specific medical fields, showing poor representation of Cochrane review authors from LMICs in the disciplines of hematology (9), gastroenterology (11), and cardiology (13). Additionally, just 12% of Cochrane’s 111,000 members were based in an LMIC in 2022 (6). Low representation of authors from LMICs may be, in part, due to the limited investment in research funding, academic institutions, and infrastructure in these countries, and in contrast, a greater relative investment in high-income countries (30).

These findings may suggest that the focus on international collaboration and standardized methodologies in Cochrane Reviews might unintentionally favour authors from high-income, English-speaking regions. The long time it takes to publish reviews could also be another barrier. New mechanisms to engage qualified researchers from a more diverse range of geographical locations to participate in Cochrane reviews might be needed. These findings may also reflect limited processes in Cochrane to ensure that individuals from LMICs are trained in Cochrane review production, and are more actively included in author teams. For example, while there are initiatives such as free access for LMIC authors to a suite of state-of-the-art online training modules, perhaps greater promotion to, and engagement with, LMIC members may help to increase author representation. Similarly, initiatives such as Cochrane Exchange (formerly Task Exchange), which are used to post review tasks to a wide audience, could potentially be used to invite authors from LMIC countries to participate in author teams more systematically, especially on topics where a global perspective.

Despite the low prevalence of LMIC-led Cochrane reviews, 22,681 Cochrane reviews included authors from 107 countries across author types, including at least 59 LMIC countries (55.1%), exhibiting significant diversity and demonstrating that academic research is indeed a global endeavour that involves contributions from scholars all across the world.

Since 1996, there has been a steady increase in female authorship in Cochrane reviews across the three author categories (first, corresponding, and last). Female first authors increased from 15.0% in 1996 to 55.6% in 2023. This is in line with research by Bhat (13) who found that the representation of females as first authors of Cochrane cardiology reviews had increased over time. In contrast to non-Cochrane reviews, the rate of growth for female first authors in Cochrane reviews is higher. There are a number of reasons that may contribute to the observed changes in gender representation. It is plausible that initiatives aimed at fostering gender equality in academia and research, an increasing appreciation of the importance of diversity and equity, and broader societal shifts toward recognizing and addressing gender disparities in various professional fields have played a role in encouraging and supporting female researchers to assume authorship roles. The greater proportions in 2023, and higher growth rate over time, for female first authors of Cochrane reviews relative to non-Cochrane reviews suggest dedicated initiatives on this front within the organisation. However, the rate of growth for female last authors in Cochrane reviews is evidently lower, representing about a third of last authors in 2023, compared to approximately half of first authors the same year. Further, while 33 of 52 CRGs had more than 50% of reviews authored by female first authors, a majority of CRGs had 25-50% female last authors (n=38), similarly exhibiting a ceiling effect for this author category. This is notable given that last authors typically represent senior positions on a review team, and may suggest that additional factors or barriers may influence the advancement of women into higher-ranking authorship roles within Cochrane.

Our findings also suggested varying diversity between Cochrane Review Groups. In the future, it would be interesting to explore how Cochrane Review Groups achieve diversity in authorship and encourage shared learning between groups, particularly within the changing landscape of Cochrane review production. The number of Cochrane Review Groups has reduced since 2023 when the National Institute for Health and Care Research ceased funding for the Cochrane Review Groups that were based in the United Kingdom (31). At the same time, seven “Cochrane Thematic Groups” were created, each focusing on an aspect of healthcare (32). Cochrane also introduced “Evidence Synthesis Units” which focus on developing locally and regionally relevant evidence and responding to diverse stakeholder needs (33). It is important that the remaining Cochrane Review Groups, and the new thematic groups, prioritize diversity and inclusion in review production.

### Implications

The findings of this study have potential implications for Cochrane review production. It is clear that Cochrane needs to do more to improve the inclusion of individuals from LMICs. Supporting individuals from LMICs as authors of Cochrane reviews will encourage varying perspectives, interests, and priorities. This is likely to lead to a wider coverage of health topics, a stronger focus on health equity, and attention to conditions with a high global burden of disease. In turn, this will help to ensure that harder-to-reach groups within the population benefit from Cochrane evidence and that intervention-generated inequalities are avoided (34). Also, it is shown that higher authorship of underrepresented groups in Cochrane reviews is associated with greater considerations of equity-related analyses in the reviews (e.g., females (35)). The Cochrane Health Equity Thematic Group is well positioned to help in this effort, as the Group aims to promote health equity within Cochrane, by supporting CRGs and author teams to consider health equity in their work, and by developing and evaluating methods to analyze health equity in systematic reviews and the evidence base (36). Working together with other organizations globally will also be crucial to improving the inclusion of people from LMICs. An example of such collaboration is the Global Evidence Synthesis Initiative (37).

The finding that there is a gender disparity in leadership roles in Cochrane reviews suggests that Cochrane would benefit from exploring ways to support female authors into senior author roles. Future research should explore potential challenges or biases that may hinder the progression of female researchers. Identifying and addressing these barriers, which could encompass institutional practices, and biases in funding and mentorship opportunities (38), is crucial for achieving a more equitable distribution of authorship responsibilities. In 2022, the U.S. National Institutes of Health developed new initiatives to promote gender equity. For example, they offer additional financial support to assist researchers in maintaining their work during childbirth, adoption, and primary caregiving duties. Additionally, they are acknowledging institutions that effectively tackle gender diversity and equity concerns, thereby promoting the adoption of proven, replicable strategies for enhancing faculty diversity (39).

Regarding geographical diversity, further research investigating citation metrics, collaboration patterns, and the significance of the research may provide a more comprehensive understanding of the impact of Cochrane reviews, and of the make-up of the entirety of author teams.

### Strengths and limitations

Strengths of this study include that it followed a pre-registered protocol and used a fully reproducible methodology to systematically extract and analyze data from Cochrane reviews. Data was extracted from Cochrane reviews using an automated technique, allowing for the collection of a large amount of data. Additionally, the study was conducted by a diverse and international team of researchers with varying backgrounds in healthcare.

Limitations include that we were unable to identify the country for 318 affiliations due to insufficient information on the website. This could be an area for improvement in data collection or reporting standards. Additionally, as the variable gender was inputted with the use of the World Gender Name Dictionary (WGND) 2, there is room for error in classification. Even though this dictionary includes an extensive list of names from many languages, our variable gender is a probabilistic proxy. However, we believe that even if we had the gender ground truth, our results would not change significantly given two reasons: (1) diversity increases in other variables, thus is likely to have an increase also in gender; and (2) the WGND 2 usability, thus is the closest that we have to ground truth and it has been used in research elsewhere, providing a powerful tool with no systematic biases. Additionally, this tool has been used in other studies (40–45). The use of an automated process to collect data also has potential limitations. For example, data cleaning is a complex procedure and prone to errors if not tested adequately. However, this was a pragmatic approach and allowed for the collection of a large amount of data that would have otherwise been impossible with the available resources.

## Conclusions

Our analysis of Cochrane Reviews revealed progress in gender diversity, with a significant increase in female first authors. However, geographic diversity remains limited, with an overrepresentation of authors from high-income, English-speaking countries. Notably, diversity varied across Review Groups, with Sexually Transmitted Infections exhibiting the highest representation from non-English speaking and low/middle-income countries. While non-Cochrane reviews showed a similar trend of increasing diversity, no causal relationship between Cochrane and non-Cochrane review trends was observed. These findings suggest that while progress has been made in gender representation, further efforts are needed to enhance geographic diversity within Cochrane Reviews. Strategies such as fostering international collaborations and exploring alternative authorship models could be implemented to achieve this goal.

## Author contributions

Conceptualization: ASM, EV, SCO, ABP; Data curation: ASM; Formal analysis: ASM, JS, ET; Investigation: ASM, EV, ABP, EN, AS, LEM; Methodology: ASM, JS, EV, SK, SCO; Project administration: ASM; Resources: ASM; Software: ASM; Supervision: ASM; Validation: ASM, SK; Visualization: ASM, JS, LM; Roles/Writing - original draft: ASM, JS, EV, ET, ABP, SK, EN, AS, LEM; ACBJ, SCO and Writing - review & editing: ASM, JS, EV, ET, ABP, SK, EN, AS, LEM; ACBJ, SCO, VAW, LM, PT.

## Data Availability

All datasets and codes of workflows used in this study are publically available (OSF: https://osf.io/fv5ys, GitHub: https://github.com/choxos/cochraneauthors).

## Notes

### Summary of Updates

We added some new results and revised the Discussion.

## References

1. Cochrane. About us [Internet]. [cited 2024 Feb 27]. Available from: https://www.cochrane.org/about-us

2. Thomas J, Kneale D, McKenzie JE, Brennan SE, Bhaumik S. Determining the scope of the review and the questions it will address. Cochrane Handb Syst Rev Interv. 2019;13–31.

3. Cochrane. Geographic Groups [Internet]. [cited 2024 Feb 27]. Available from: https://www.cochrane.org/about-us/our-global-community/geographic-groups

4. Cochrane. Strategy for Change [Internet]. [cited 2024 Feb 27]. Available from: https://www.cochrane.org/about-us/strategy-for-change

5. Cochrane. Cochrane Membership [Internet]. [cited 2024 Mar 23]. Available from: https://www.cochrane.org/join-cochrane/membership

6. Cochrane. How could Cochrane be even more inclusive? Feedback from over 1300 people [Internet]. Cochrane; 2022 [cited 2024 Feb 27]. Available from: https://www.cochrane.org/sites/default/files/public/uploads/pdf/cochrane-listen-and-learn-diversity.pdf

7. Cochrane. Current Cochrane Group Priority Setting Projects [Internet]. [cited 2024 Feb 27]. Available from: https://community.cochrane.org/news/current-cochrane-group-priority-setting-projects

8. Tomlinson E, Pardo Pardo J, Sivesind T, Szeto MD, Laughter M, Foxlee R, et al. Prioritising Cochrane reviews to be updated with health equity focus. Int J Equity Health. 2023 May 5;22(1):81.

9. Biswas J, Dhali A, Rathna RB, D’Souza C. Authorship diversity in hematologyl_Jrelated Cochrane systematic reviews: Inequities in global representation. Res Pract Thromb Haemost. 2022 Aug;6(6):e12778.

10. Qureshi R, Han G, Fapohunda K, Abariga S, Wilson R, Li T. Authorship diversity among systematic reviews in eyes and vision. Syst Rev. 2020 Dec;9(1):192.

11. Dhali A, D’Souza C, Rathna RB, Biswas J, Dhali GK. Authorship diversity in Gastroenterology-related Cochrane systematic reviews: Inequities in global representation. Front Med. 2022 Sep 2;9:982664.

12. Evans J, Mwangi N, Burn H, Ramke J. Equity was rarely considered in Cochrane Eyes and Vision systematic reviews and primary studies on cataract. J Clin Epidemiol. 2020 Sep;125:57–63.

13. Bhat V, Ozair A, Bellur S, Subhash NR, Kumar A, Majmundar M, et al. Inequities in Country- and Gender-Based Authorship Representation in Cardiology-Related Cochrane Reviews. JACC Adv. 2022 Dec;1(5):100140.

14. Rathna RB, Biswas J, D’Souza C, Joseph JM, Kipkorir V, Dhali A. Authorship diversity in general surgery-related Cochrane systematic reviews: a bibliometric study. Br J Surg. 2023 Jul 17;110(8):989–90.

15. Tomlinson E, Petkovic J, Welch V, Tugwell P. It’s time to increase the global relevance of Cochrane Reviews by applying an ‘equity lens.’ Cochrane Editorial Unit, editor. Cochrane Database Syst Rev [Internet]. 2021 Oct 28 [cited 2023 Dec 4];2021(10). Available from: http://doi.wiley.com/10.1002/14651858.ED000155

16. O’Neill J, Tabish H, Welch V, Petticrew M, Pottie K, Clarke M, et al. Applying an equity lens to interventions: using PROGRESS ensures consideration of socially stratifying factors to illuminate inequities in health. J Clin Epidemiol. 2014 Jan;67(1):56–64.

17. Mbuagbaw L, Schoonees A, Oliver J, Arikpo D, Durão S, Effa E, et al. Publication practices of sub-Saharan African Cochrane authors: a bibliometric study. BMJ Open. 2021 Sep;11(9):e051839.

18. Martínez GL, de Juano-i-Ribes HS, Yin D, Le Feuvre B, Hamdan-Livramento I, Saito K, et al. Expanding the World Gender-Name Dictionary: WGND 2.0. World Intellectual Property Organization-Economics and Statistics Division; 2021.

19. Ameratunga D, Yazdani A, Kroon B. Antibiotics prior to or at the time of embryo transfer in ART. Cochrane Gynaecology and Fertility Group, editor. Cochrane Database Syst Rev [Internet]. 2023 Nov 23 [cited 2023 Dec 2];2023(11). Available from: http://doi.wiley.com/10.1002/14651858.CD008995.pub3

20. Cundy O, Lange CA, Bunce C, Bainbridge JW, Solebo AL. Face-down positioning or posturing after macular hole surgery. Cochrane Eyes and Vision Group, editor. Cochrane Database Syst Rev [Internet]. 2023 Nov 21 [cited 2023 Dec 2];2023(11). Available from: http://doi.wiley.com/10.1002/14651858.CD008228.pub3

21. Heneghan M, Southern KW, Murphy J, Sinha IP, Nevitt SJ. Corrector therapies (with or without potentiators) for people with cystic fibrosis with class II CFTR gene variants (most commonly F508del). Cochrane Cystic Fibrosis and Genetic Disorders Group, editor. Cochrane Database Syst Rev [Internet]. 2023 Nov 20 [cited 2023 Dec 2];2023(11). Available from: http://doi.wiley.com/10.1002/14651858.CD010966.pub4

22. Roberts L, Lin L, Alsweiler J, Edwards T, Liu G, Harding JE. Oral dextrose gel to prevent hypoglycaemia in at-risk neonates. Cochrane Neonatal Group, editor. Cochrane Database Syst Rev [Internet]. 2023 Nov 28 [cited 2023 Dec 2];2023(11). Available from: http://doi.wiley.com/10.1002/14651858.CD012152.pub4

23. Tunnicliffe DJ, Palmer SC, Cashmore BA, Saglimbene VM, Krishnasamy R, Lambert K, et al. HMG CoA reductase inhibitors (statins) for people with chronic kidney disease not requiring dialysis. Cochrane Kidney and Transplant Group, editor. Cochrane Database Syst Rev [Internet]. 2023 Nov 29 [cited 2023 Dec 2];2023(11). Available from: http://doi.wiley.com/10.1002/14651858.CD007784.pub3

24. Sayers E. E-utilities Quick Start. In: Entrez Programming Utilities Help [Internet]. Bethesda (MD): National Center for Biotechnology Information (US); 2008 [cited 2024 Feb 27]. Available from: https://www.ncbi.nlm.nih.gov/books/NBK25500/

25. R Core Team. R: A Language and Environment for Statistical Computing [Internet]. Vienna, Austria: R Foundation for Statistical Computing; 2023. Available from: https://www.R-project.org/

26. Wickham H. rvest: Easily Harvest (Scrape) Web Pages [Internet]. 2022. Available from: https://CRAN.R-project.org/package=rvest

27. Wickham H. ggplot2: Elegant Graphics for Data Analysis. 2016; Available from: https://ggplot2.tidyverse.org

28. Granger CWJ. Investigating Causal Relations by Econometric Models and Cross-spectral Methods. Econometrica. 1969 Aug;37(3):424.

29. Hiemstra C, Jones JD. Testing for Linear and Nonlinear Granger Causality in the Stock Pricel_JVolume Relation. J Finance. 1994 Dec;49(5):1639–64.

30. Franzen SRP, Chandler C, Lang T. Health research capacity development in low and middle income countries: reality or rhetoric? A systematic meta-narrative review of the qualitative literature. BMJ Open. 2017 Jan 27;7(1):e012332.

31. Pearson H. Medical-evidence giant Cochrane battles funding cuts and closures. Nature. 2023 Sep 7;621(7977):13–4.

32. Cochrane. Major milestones and new beginnings: Cochrane announces first round of new Thematic Groups [Internet]. [cited 2024 Mar 23]. Available from: https://community.cochrane.org/news/major-milestones-and-new-beginnings-cochrane-announces-first-round-new-thematic-groups

33. Cochrane. About Cochrane’s new production model [Internet]. [cited 2024 Mar 23]. Available from: https://community.cochrane.org/organizational-info/plans/future-evidence-synthesis-cochrane/about-cochranes-new-production-model

34. Welch VA, Petkovic J, Jull J, Hartling L, Klassen T, Kristjansson E, et al. Equity and specific populations. In: Cochrane Handbook for Systematic Reviews of Interventions. Wiley Online Library; 2019. p. 433–49.

35. Antequera A, Cuadrado-Conde MA, Roy-Vallejo E, Montoya-Martínez M, León-García M, Madrid-Pascual O, et al. Lack of sex-related analysis and reporting in Cochrane Reviews: a cross-sectional study. Syst Rev. 2022 Dec 26;11(1):281.

36. Parker R, Petkovic J, Pardo Pardo J, Darzi A, Dewidar O, Khabsa J, et al. The equity group: Supporting Cochrane’s social responsibility of improving health equity. Cochrane Evid Synth Methods. 2023;1(3):e12012.

37. Cochrane. Beirut - the new home of Global Evidence Synthesis Initiative [Internet]. [cited 2024 Mar 23]. Available from: https://www.cochrane.org/news/beirut-new-home-global-evidence-synthesis-initiative

38. Association of American Medical Colleges. The State of Women in Academic Medicine 2018-2019: Exploring Pathways to Equity [Internet]. Association of American Medical Colleges; 2020. Available from: https://store.aamc.org/the-state-of-women-in-academic-medicine-2018-2019-exploring-pathways-to-equity.html

39. Ten Hagen KG, Wolinetz C, Clayton JA, Bernard MA. Community voices: NIH working toward inclusive excellence by promoting and supporting women in science. Nat Commun. 2022 Mar 25;13(1):1682.

40. Bhagat V. Data and Techniques Used for Analysis of Women Authorship in STEMM: A Review. Fem Res. 2019 Oct 2;2(2):76–87.

41. Forkin KT, Render CM, Staffa SJ, Goobie SM. Trends in Gender of Authors of Patient Blood Management Publications. Anesth Analg [Internet]. 2023 Dec 28 [cited 2024 Mar 23]; Available from: https://journals.lww.com/10.1213/ANE.0000000000006749

42. Chien CV, Ouellette LL. Improving equity in patent inventorship. Science. 2023 Dec 8;382(6675):1128–9.

43. Hattke F, Vogel R. Theories and theorizing in public administration: A systematic review. Public Adm Rev. 2023 Nov;83(6):1542–63.

44. Lax-Martinez G, Raffo JD, Saito K. Identifying the gender of PCT inventors. SSRN Electron J [Internet]. 2023 [cited 2024 Mar 23]; Available from: https://www.ssrn.com/abstract=4434107

45. Keenan P, Heavin C. DSS research: a bibliometric analysis by gender. J Decis Syst. 2022 Dec 15;31(sup1):107–16.

