## Appendix 1 for "Geographical and Gender Diversity in Cochrane and non-Cochrane Reviews Authorship: A Meta-Research Study"

**Appendix 1.** Deviations from the protocol.

1. In the protocol, we aimed to analyze only the latest version of the reviews. We changed the inclusion criteria to all the published papers.
2. Also, we included analyses for Spanish-speaking countries.
3. Another change was switching from non-high-income OECD to lower-and-middle-income countries (LMICs). We chose the former since the number of authors from high-income non-OECD countries was rare. However, to be more concordant with the publications in the fields, we considered LMICs.
4. Comparison with non-Cochrane reviews was not forecasted in the protocol. During the analysis phase, we were informed about the ability to assess the non-Cochrane reviews automatically. Since this added extensive value to our study, we added this to our aims.
