## Appendix 2 for "Geographical and Gender Diversity in Cochrane and non-Cochrane Reviews Authorship: A Meta-Research Study"

Ahmad Sofi-Mahmudi

### 1 Aim

We aimed to determine the level of country, region, language, and gender diversity in Cochrane Reviews' authorship.

#### 2 Results

First, loading the needed packages:

```
pacman::p_load(knitr,
               lubridate,
               ggplot2,
               dplyr,
               tidyr,
               stringr,
               lme4,
               openxlsx,
               sf,
               lme4,
               lmtest,
               ggpubr)
```

And then, loading the datasets:

```
cochrane = read.csv("data/cochraneauthors_db_final.csv")
```

##### 2.1 General characteristics

The number of citations retrieved from the Cochrane Library on November 6, 2023 was 9,153. Adding previous versions of the reviews to the database increased the sample size to 22,681 articles:

```
nrow(cochrane)
```

```
[1] 22681
```

The highest number of authors belonged to two reviews with 41 authors by Philipp Schuetz and Sarah Burdett from Acute Respiratory Infections and Lung Cancer Groups:

```
kable(cochrane[which(is.na(cochrane$aff41) == F), c(2,5,1,6)])
```

| | X.1 | date | X.2 | type |
|---|---|---|---|---|
| 14453 | 14453 | 2017-10-12 | 14453 | Intervention |

| | X.1 | date | X.2 | type |
|---|---|---|---|---|
| 22318 | 22318 | 2015-03-02 | 22318 | Intervention |

Now, let's take a look at the publication years:

```
cochrane = cochrane %>% mutate(year = year(date))

kable(cochrane %>%
  group_by(year) %>%
  summarize(count = n()) %>%
  mutate(percentage = round(count / sum(count) * 100, 2)))
```

| year | count | percentage |
|---|---|---|
| 1995 | 1 | 0.00 |
| 1996 | 60 | 0.26 |
| 1997 | 47 | 0.21 |
| 1998 | 131 | 0.58 |
| 1999 | 207 | 0.91 |
| 2000 | 345 | 1.52 |
| 2001 | 531 | 2.34 |
| 2002 | 666 | 2.94 |
| 2003 | 864 | 3.81 |
| 2004 | 838 | 3.69 |
| 2005 | 868 | 3.83 |
| 2006 | 959 | 4.23 |
| 2007 | 1038 | 4.58 |
| 2008 | 959 | 4.23 |
| 2009 | 1241 | 5.47 |
| 2010 | 1231 | 5.43 |
| 2011 | 1174 | 5.18 |
| 2012 | 1508 | 6.65 |
| 2013 | 1457 | 6.42 |
| 2014 | 1284 | 5.66 |
| 2015 | 1381 | 6.09 |
| 2016 | 1145 | 5.05 |
| 2017 | 1050 | 4.63 |
| 2018 | 856 | 3.77 |
| 2019 | 741 | 3.27 |
| 2020 | 731 | 3.22 |
| 2021 | 619 | 2.73 |

| year | count | percentage |
|---|---|---|
| 2022 | 373 | 1.64 |
| 2023 | 376 | 1.66 |

It's always good to have a graphical look as well:

```
cochrane %>% ggplot() + aes(x = year) +
  geom_bar(fill="steelblue") +
  labs(title = "Number of Cochrane reviews in each year",
       x = "Year",
       y = "Count") +
  theme(axis.text.x = element_text(angle = 45, vjust = 0.5, hjust=1)) +
  theme_minimal()
```

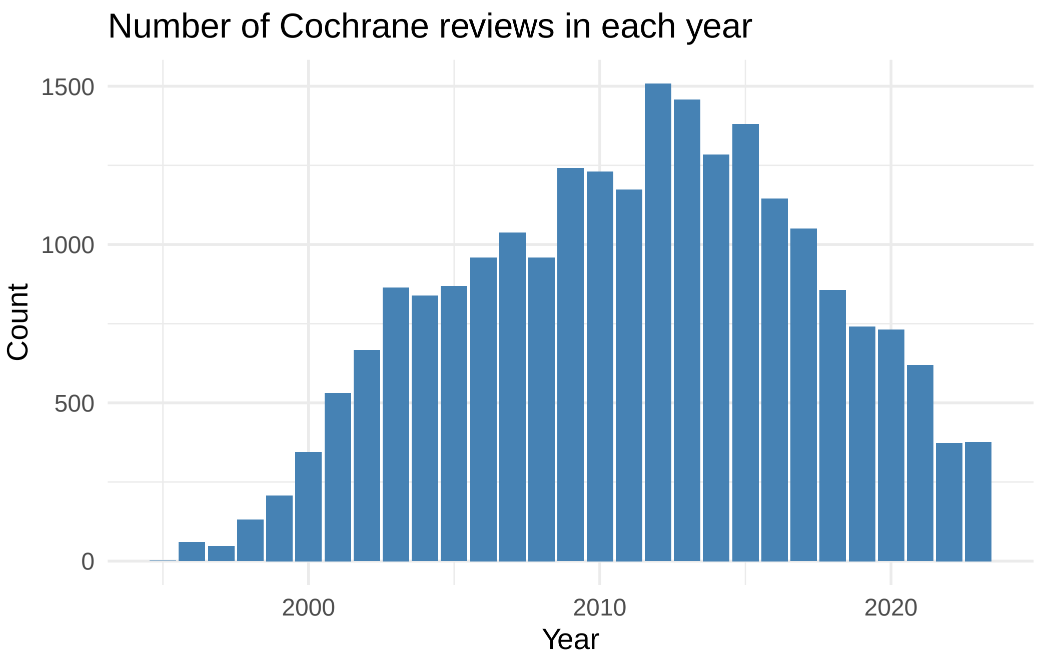

```
# ggsave("figures/Fig1.png", width = 10, height = 5, dpi = 300)
```

Review types:

```
kable(cochrane %>%
  group_by(type) %>%
  summarize(count = n()) %>%
```

```
  mutate(percentage = round(count / sum(count) * 100, 2)) %>%
  arrange(desc(count)))
```

| type | count | percentage |
|---|---|---|
| Intervention | 21965 | 96.84 |
| Diagnostic | 358 | 1.58 |
| Overview | 140 | 0.62 |
| Methodology | 110 | 0.48 |
| Qualitative | 44 | 0.19 |
| Prognosis | 36 | 0.16 |
| Rapid | 15 | 0.07 |
| Prototype | 13 | 0.06 |

Review stages:

```
kable(cochrane %>%
  group_by(stage) %>%
  summarize(count = n()) %>%
  mutate(percentage = round(count / sum(count) * 100, 2)) %>%
  arrange(desc(count)))
```

| stage | count | percentage |
|---|---|---|
| Review | 15524 | 68.44 |
| Protocol | 7157 | 31.56 |

Let's see how many article each review group has produced:

```
kable(cochrane %>%
  group_by(group) %>%
  summarize(count = n()) %>%
  mutate(percentage = round(count / sum(count) * 100, 2)) %>%
  arrange(desc(count)))
```

| group | count | percentage |
|---|---|---|
| Cochrane Pregnancy and Childbirth Group | 1634 | 7.20 |
| Cochrane Neonatal Group | 1118 | 4.93 |

| group | count | percentage |
|---|---|---|
| Cochrane Airways Group | 873 | 3.85 |
| Cochrane Pain, Palliative and Supportive Care Group | 785 | 3.46 |
| Cochrane Gynaecology and Fertility Group | 765 | 3.37 |
| Cochrane Cystic Fibrosis and Genetic Disorders Group | 750 | 3.31 |
| Cochrane Eyes and Vision Group | 672 | 2.96 |
| Cochrane Acute Respiratory Infections Group | 629 | 2.77 |
| Cochrane Gut Group | 628 | 2.77 |
| Cochrane Gynaecological, Neuro-oncology and Orphan Cancer Group | 604 | 2.66 |
| Cochrane Oral Health Group | 588 | 2.59 |
| Cochrane Heart Group | 586 | 2.58 |
| Cochrane Hepato-Biliary Group | 580 | 2.56 |
| Cochrane Stroke Group | 577 | 2.54 |
| Cochrane Vascular Group | 563 | 2.48 |
| Cochrane Kidney and Transplant Group | 557 | 2.46 |
| Cochrane Schizophrenia Group | 556 | 2.45 |
| Cochrane Anaesthesia Group | 493 | 2.17 |
| Cochrane Infectious Diseases Group | 484 | 2.13 |
| Cochrane Common Mental Disorders Group | 480 | 2.12 |
| Cochrane Wounds Group | 477 | 2.10 |
| Cochrane Developmental, Psychosocial and Learning Problems Group | 474 | 2.09 |
| Cochrane Musculoskeletal Group | 463 | 2.04 |
| Cochrane Neuromuscular Group | 426 | 1.88 |
| Cochrane Effective Practice and Organisation of Care Group | 423 | 1.86 |
| Cochrane Dementia and Cognitive Improvement Group | 414 | 1.83 |
| Cochrane Bone, Joint and Muscle Trauma Group | 400 | 1.76 |
| Cochrane ENT Group | 385 | 1.70 |
| Cochrane Injuries Group | 371 | 1.64 |
| Cochrane Epilepsy Group | 361 | 1.59 |
| Cochrane Metabolic and Endocrine Disorders Group | 326 | 1.44 |
| Cochrane Colorectal Group | 312 | 1.38 |
| Cochrane Tobacco Addiction Group | 299 | 1.32 |
| Cochrane Skin Group | 298 | 1.31 |
| Cochrane Incontinence Group | 266 | 1.17 |
| Cochrane Drugs and Alcohol Group | 259 | 1.14 |
| Cochrane Fertility Regulation Group | 250 | 1.10 |
| Cochrane HIV/AIDS Group | 231 | 1.02 |
| Cochrane Back and Neck Group | 224 | 0.99 |
| Cochrane Haematology Group | 219 | 0.97 |
| Cochrane Hypertension Group | 211 | 0.93 |
| Cochrane Breast Cancer Group | 208 | 0.92 |
| Cochrane Urology Group | 204 | 0.90 |

| group | count | percentage |
|---|---|---|
| Cochrane Consumers and Communication Group | 182 | 0.80 |
| Cochrane Multiple Sclerosis and Rare Diseases of the CNS Group | 155 | 0.68 |
| Cochrane Movement Disorders Group | 149 | 0.66 |
| Cochrane Emergency and Critical Care Group | 139 | 0.61 |
| Cochrane Public Health Group | 124 | 0.55 |
| Cochrane Childhood Cancer Group | 118 | 0.52 |
| Cochrane Lung Cancer Group | 109 | 0.48 |
| Cochrane Work Group | 108 | 0.48 |
| Cochrane Methodology Group | 104 | 0.46 |
| Cochrane Sexually Transmitted Infections Group | 47 | 0.21 |
| Cochrane ENT GroupCochrane Oral Health Group | 6 | 0.03 |
| Cochrane Infectious Diseases GroupCochrane Haematology Group | 4 | 0.02 |
| Cochrane Back and Neck GroupCochrane Musculoskeletal Group | 3 | 0.01 |
| Cochrane Heart GroupCochrane Stroke Group | 2 | 0.01 |
| Cochrane Methodology GroupCochrane Effective Practice and Organisation of Care Group | 2 | 0.01 |
| Cochrane Common Mental Disorders GroupCochrane Developmental, Psychosocial and Learning Problems Group | 1 | 0.00 |
| Cochrane Gynaecological, Neuro-oncology and Orphan Cancer GroupCochrane Childhood Cancer Group | 1 | 0.00 |
| Cochrane Gynaecological, Neuro-oncology and Orphan Cancer GroupCochrane Haematology Group | 1 | 0.00 |
| Cochrane Gynaecology and Fertility GroupCochrane Incontinence Group | 1 | 0.00 |
| Cochrane Heart GroupCochrane Vascular Group | 1 | 0.00 |
| Cochrane Public Health GroupCochrane Heart Group | 1 | 0.00 |

##### 2.1.1 Yearly trend by review group

Since some reviews were a collaboration between two review groups, we add a new column and only include those reviews which were from just one review group:

```
cochrane = cochrane %>%
  mutate(group_one = ifelse(str_count(group, "Cochrane") == 1, group, NA))
```

Then,

```
cochrane %>%
  group_by(year, group_one) %>%
  filter(is.na(group_one) == F) %>%
```

```
  summarise(n = n()) %>%
  ggplot() +
  aes(x = year, y = n) +
  geom_line() +
  labs(title = "Number of Cochrane reviews in each year",
       x = "Year",
       y = "Count") +
  theme(axis.text.x = element_text(angle = 45, vjust = 0.5, hjust=1)) +
  facet_wrap(~group_one, ncol = 10)
```

```
`summarise()` has grouped output by 'year'. You can override using the
`.groups` argument.
```

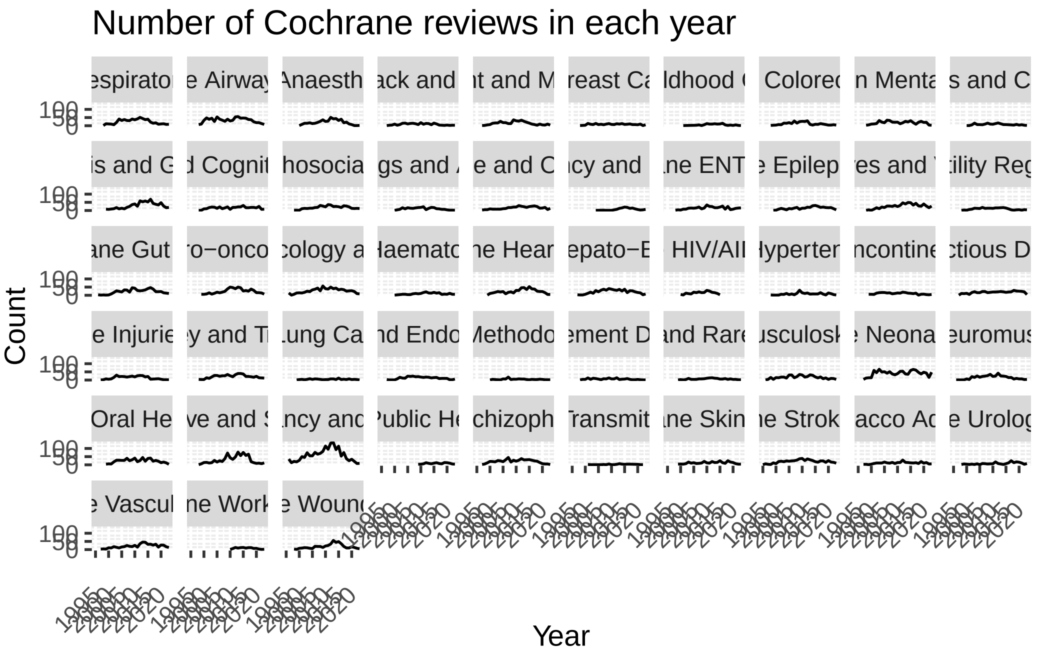

```
# ggsave("figures/Appendix2.png", width = 20, height = 12, dpi = 300)
```

##### 2.2 Geographical diversity

Since the year 1995 has only 1 entry in our dataset, we drop it:

```
cochrane = cochrane %>% filter(year != 1995)
```

The number of countries detected successfully:

```
cochrane$first_author_country[cochrane$first_author_country == ""] <- NA

sum(table(cochrane$first_author_country))
```

```
[1] 22363
```

##### 2.2.1 First authors

The overall number of countries:

```
length(unique(cochrane$first_author_country))
```

```
[1] 102
```

Countries with the highest number of first authors:

```
top_first = cochrane %>%
  filter(!is.na(first_author_country)) %>%
  group_by(first_author_country) %>%
  summarize(count = n()) %>%
  mutate(percentage = round(count / sum(count) * 100, 2)) %>%
  arrange(desc(count))

names(top_first)[1] = c("country")

kable(top_first[1:10,])
```

| country | count | percentage |
|---|---:|---:|
| United Kingdom | 7426 | 33.21 |
| Australia | 2595 | 11.60 |
| United States | 1559 | 6.97 |
| Canada | 1475 | 6.60 |
| China | 1048 | 4.69 |
| Netherlands | 1005 | 4.49 |
| Germany | 652 | 2.92 |
| Italy | 566 | 2.53 |
| Brazil | 565 | 2.53 |

| country | count | percentage |
|---|---|---|
| New Zealand | 450 | 2.01 |

World heat map:

```
# Loading the world map
world_map = map_data("world")
world_map = subset(world_map, region != "Antarctica")

# Some modifications are needed
top_first$country <- as.character(top_first$country)
top_first$country[top_first$country == "United States"] <- "USA"
top_first$country[top_first$country == "United Kingdom"] <- "UK"
top_first$country = as.factor(top_first$country)

# Drawing the map
top_first_map = ggplot(top_first) +
  geom_map(
    dat = world_map, map = world_map, aes(map_id = region),
    fill = "white", color = "#7f7f7f", size = 0.25
  ) +
  geom_map(map = world_map, aes(map_id = country, fill = log10(count)), size = 0.25) +
  scale_fill_gradient(low = "snow2", high = "red2", name = "Total first authors (log10 scale
  expand_limits(x = world_map$long, y = world_map$lat) + theme(legend.position="bottom",
        axis.line=element_blank(),
        axis.text=element_blank(),
        axis.ticks=element_blank(),
        axis.title=element_blank(),
        panel.background=element_blank(),
        panel.border=element_blank(),
        panel.grid=element_blank())
```

```
Warning: Using `size` aesthetic for lines was deprecated in ggplot2 3.4.0.
i Please use `linewidth` instead.
```

```
top_first_map
```

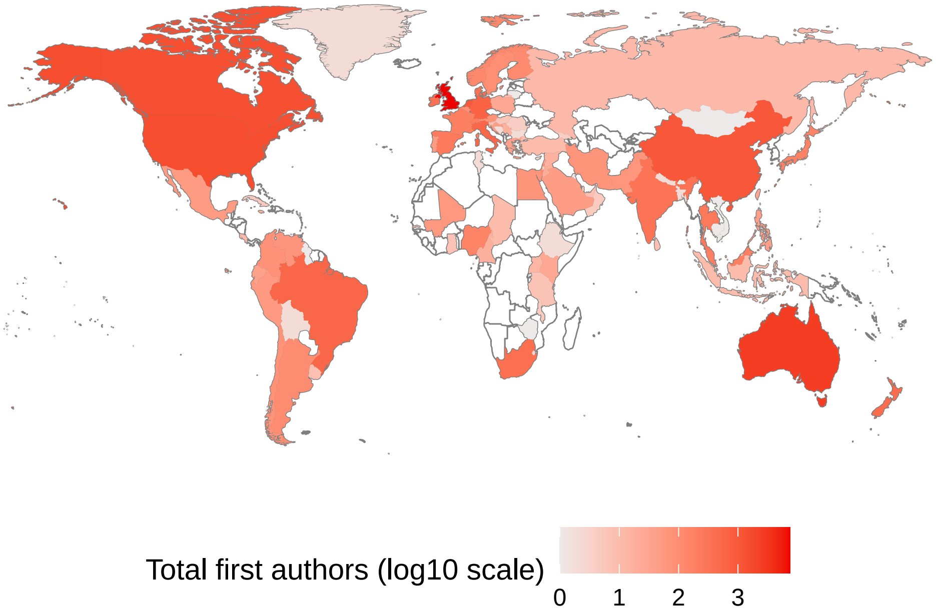

```r
# ggsave("figures/Fig2A.png", width = 10, height = 5, dpi = 300)
```

###### 2.2.1.1 World Bank Income Regions

```r
kable(cochrane %>%
  filter(!is.na(first_author_region_income)) %>%
  group_by(first_author_region_income) %>%
  summarize(count = n()) %>%
  mutate(percentage = round(count / sum(count) * 100, 2)) %>%
  arrange(desc(count)))
```

| first_author_region_income | count | percentage |
|---|---|---|
| 1. High income: OECD | 18049 | 81.26 |
| 3. Upper middle income | 3146 | 14.16 |
| 4. Lower middle income | 729 | 3.28 |
| 2. High income: nonOECD | 146 | 0.66 |
| 5. Low income | 141 | 0.63 |

High income: OECD vs. non-High income: OECD (based on the WB income regions):

First, making new columns for High income: OECD vs. non-High income: OECD

```
cochrane = cochrane %>% mutate(first_author_lowincome = ifelse(first_author_region_income ==
                    corresponding_author_lowincome = ifelse(corresponding_author_region_incon
                    last_author_lowincome = ifelse(last_author_region_income == "1. High inc
```

Then, making a line graph for all three indicators:

```
yearly_trend_region_lowincome =
        cochrane %>%
        select(year,
               first_author_lowincome,
               corresponding_author_lowincome,
               last_author_lowincome) %>%
        gather("indicator", "value", -year) %>%
        count(year, indicator, value) %>%
        mutate(indicator = recode(indicator,
                                  first_author_lowincome = "First authors from non-High-incon
                                  corresponding_author_lowincome = "Corresponding authors fro
                                  last_author_lowincome = "Last authors from non-High-income
        complete(indicator, value, year, fill = list(n = 0)) %>%
        group_by(year, indicator) %>%
        mutate(p = n / sum(n)) %>%
        filter(value) %>%
        ungroup()

plot_trend_region_lowincome =
        yearly_trend_region_lowincome %>%
        ggplot() +
        aes(x = year,
            y = p,
            group = indicator,
            color = indicator) +
        geom_line(size = 0.75) +
        scale_y_continuous(limits = c(0, 1),
                           labels = scales::percent) +
        scale_color_discrete(name = NULL) +
        scale_fill_discrete(breaks = c("First authors from non-High-income OECD countries",
                                       "Corresponding authors from non-High-income OECD coun
                                       "Last authors from non-High-income OECD countries")) +
        labs(title="Region diversity in Cochrane Reviews' authorship: non-High-income OECD vs
               y = "Proportion of authors from non-High-income OECD countries (%)",
               x = "Year") +
        theme(panel.grid.minor = element_blank(),
```

```
                legend.position = c(0.3, 0.5),
                axis.text.x = element_text(angle = 45, vjust = 0.9, hjust=1)
                )
```

```
Warning: A numeric `legend.position` argument in `theme()` was deprecated in ggplot2
3.5.0.
i Please use the `legend.position.inside` argument of `theme()` instead.
```

```
#png("figures/plot_trend_region_highincome.png", width = 10, height = 7, units = "in", res =
plot_trend_region_lowincome
```

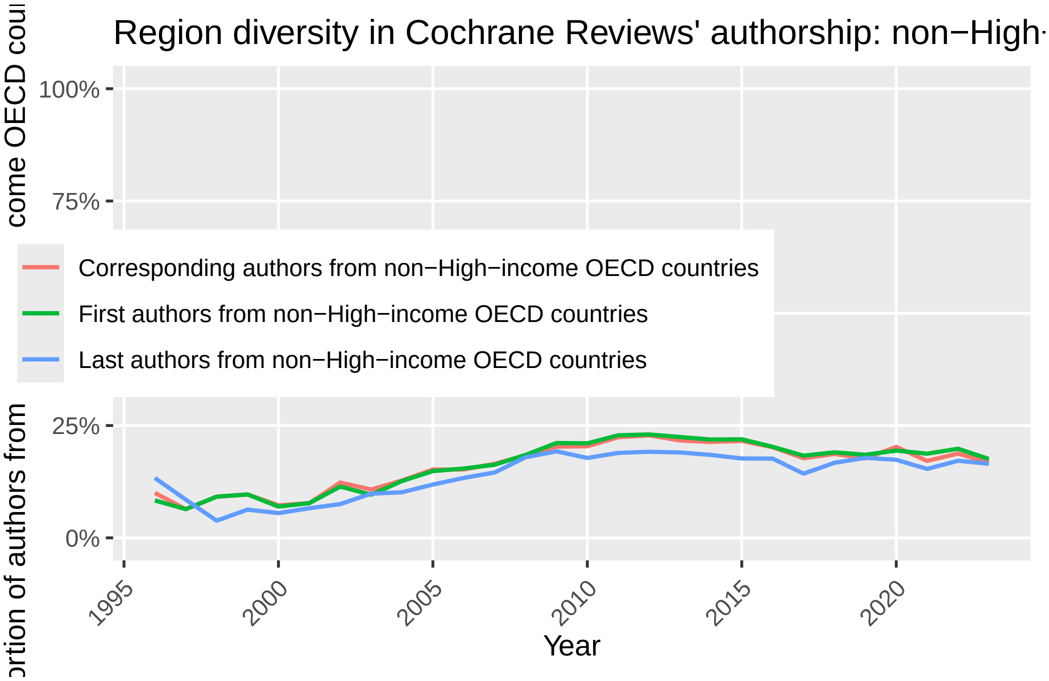

```
# ggsave("figures/Fig3.png", width = 10, height = 7, dpi = 300)
#dev.off()
```

###### 2.2.1.2 English-speaking

```
cochrane = cochrane %>% mutate(first_author_english = ifelse(is.na(first_author_country) == 

kable(cochrane %>%
  filter(!is.na(first_author_english)) %>%
```

```
  group_by(first_author_english) %>%
  summarize(count = n()) %>%
  mutate(percentage = round(count / sum(count) * 100, 2)) %>%
  arrange(desc(count))
```

| first_author_english | count | percentage |
|---|---|---|
| Yes | 13866 | 62 |
| No | 8497 | 38 |

We have the column. So, we directly make a line graph for all three indicators:

```
yearly_trend_region_non_english =
        cochrane %>%
        select(year,
               first_author_english,
               corresponding_author_english,
               last_author_english) %>%
        gather("indicator", "value", -year) %>%
        count(year, indicator, value) %>%
        mutate(indicator = recode(indicator,
                                  first_author_english = "First authors from non-English-spea[illegible]
                                  corresponding_author_english = "Corresponding authors from [illegible]
                                  last_author_english = "Last authors from non-English-speak[illegible]
        complete(indicator, value, year, fill = list(n = 0)) %>%
        group_by(year, indicator) %>%
        mutate(p = n / sum(n)) %>%
        filter(value == "No") %>%
        ungroup()

plot_trend_region_non_english =
        yearly_trend_region_non_english %>%
        ggplot() +
        aes(x = year,
            y = p,
            group = indicator,
            color = indicator) +
        geom_line(size = 0.75) +
        scale_y_continuous(limits = c(0, 1),
                           labels = scales::percent) +
        scale_color_discrete(name = NULL) +
```

```
        scale_fill_discrete(breaks = c("First authors from non-English-speaking countries",
                                       "Corresponding authors from non-English-speaking coun
                                       "Last authors from non-English-speaking countries")) 
        labs(   title="Region diversity in Cochrane Reviews' authorship: non-English-speakin
                y = "Proportion of authors from non-English-speaking countries (%)",
                x = "Year") +
        theme(panel.grid.minor = element_blank(),
              legend.position = c(0.25, 0.5),
              axis.text.x = element_text(angle = 45, vjust = 0.9, hjust=1)
              )
#png("figures/plot_trend_region_non_english.png", width = 10, height = 7, units = "in", res 
plot_trend_region_non_english
```

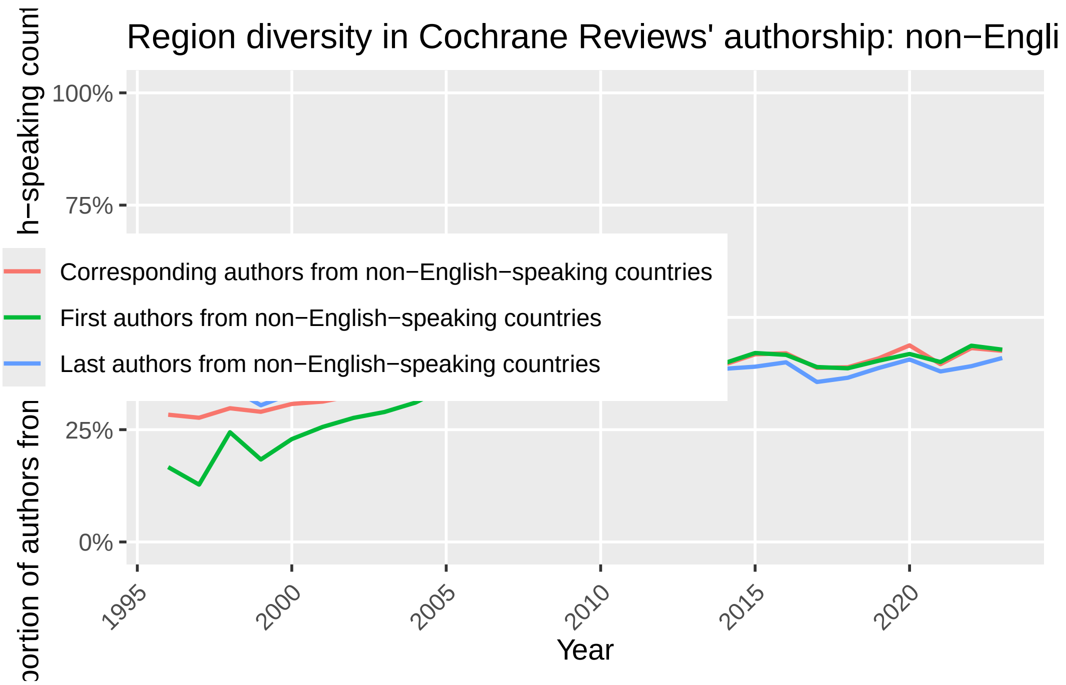

```
# ggsave("figures/Fig4.png", width = 10, height = 7, dpi = 300)
#dev.off()
```

###### 2.2.1.3 Spanish-speaking

```
cochrane = cochrane %>% mutate(first_author_spanish = ifelse(is.na(first_author_country) == 

kable(cochrane %>%
```

```
  filter(!is.na(first_author_spanish)) %>%
  group_by(first_author_spanish) %>%
  summarize(count = n()) %>%
  mutate(percentage = round(count / sum(count) * 100, 2)) %>%
  arrange(desc(count))
```

| first_author_spanish | count | percentage |
|---|---|---|
| FALSE | 21610 | 96.63 |
| TRUE | 753 | 3.37 |

We have the column. So, we directly make a line graph for all three indicators:

```
yearly_trend_region_spanish =
        cochrane %>%
        select(year,
               first_author_spanish,
               corresponding_author_spanish,
               last_author_spanish) %>%
        gather("indicator", "value", -year) %>%
        count(year, indicator, value) %>%
        mutate(indicator = recode(indicator,
                                  first_author_spanish = "First authors from Spanish-speaking
                                  corresponding_author_spanish = "Corresponding authors from
                                  last_author_spanish = "Last authors from Spanish-speaking c
        complete(indicator, value, year, fill = list(n = 0)) %>%
        group_by(year, indicator) %>%
        mutate(p = n / sum(n)) %>%
        filter(value == TRUE) %>%
        ungroup()

plot_trend_region_spanish =
        yearly_trend_region_spanish %>%
        ggplot() +
        aes(x = year,
            y = p,
            group = indicator,
            color = indicator) +
        geom_line(size = 0.75) +
        scale_y_continuous(limits = c(0, 1),
                           labels = scales::percent) +
```

```
        scale_color_discrete(name = NULL) +
        scale_fill_discrete(breaks = c("First authors from Spanish-speaking countries",
                                       "Corresponding authors from Spanish-speaking countries
                                       "Last authors from Spanish-speaking countries")) +
        labs(title="Region diversity in Cochrane Reviews' authorship: non-Spanish-speaking vs
                y = "Proportion of authors from non-Spanish-speaking countries (%)",
                x = "Year") +
        theme(panel.grid.minor = element_blank(),
              legend.position = c(0.25, 0.5),
              axis.text.x = element_text(angle = 45, vjust = 0.9, hjust=1)
              )
#png("figures/plot_trend_region_non_spanish.png", width = 10, height = 7, units = "in", res =
plot_trend_region_spanish
```

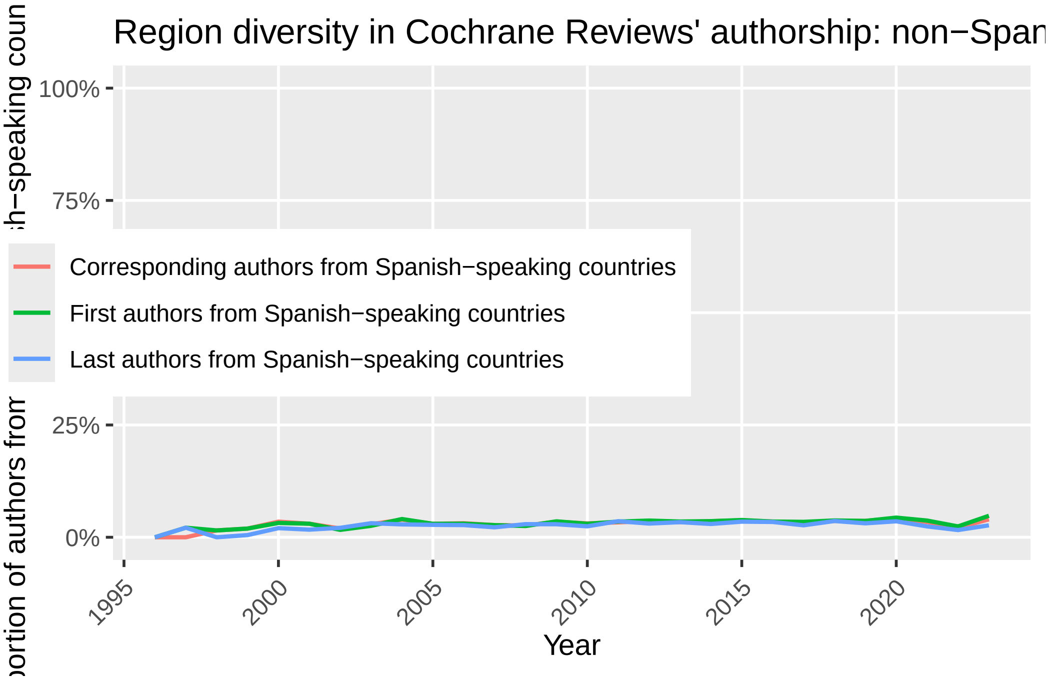

```
# ggsave("figures/Fig4.png", width = 10, height = 7, dpi = 300)
#dev.off()
```

###### 2.2.1.3.1 Combining all plots into one

```
ggarrange(plot_trend_region_lowincome, plot_trend_region_non_english, plot_trend_region_spani
                    labels = c("A", "B", "C"),
                    ncol = 1)
```

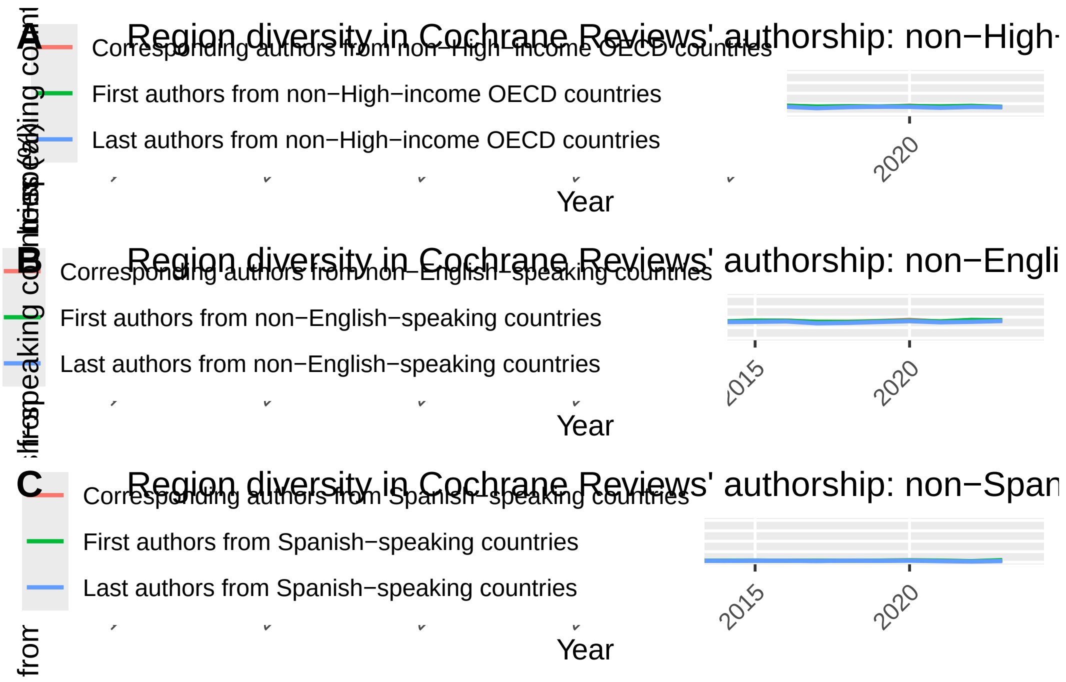

```
# ggsave("figures/Fig3.png", width = 10, height = 21, dpi = 300)
```

###### 2.2.2 Corresponding authors

The overall number of countries:

```
cochrane$corresponding_author_country[cochrane$corresponding_author_country == ""] <- NA

length(unique(cochrane$corresponding_author_country))
```

```
[1] 98
```

Countries with the highest number of corresponding authors:

```
top_corresponding = cochrane %>%
  filter(!is.na(corresponding_author_country)) %>%
  group_by(corresponding_author_country) %>%
  summarize(count = n()) %>%
  mutate(percentage = round(count / sum(count) * 100, 2)) %>%
  arrange(desc(count))
```

```r
names(top_corresponding)[1] = c("country")

kable(top_corresponding[1:10,])
```

| country | count | percentage |
|---|---|---|
| United Kingdom | 7412 | 33.18 |
| Australia | 2619 | 11.72 |
| United States | 1537 | 6.88 |
| Canada | 1516 | 6.79 |
| China | 1069 | 4.79 |
| Netherlands | 1008 | 4.51 |
| Germany | 651 | 2.91 |
| Italy | 564 | 2.52 |
| Brazil | 562 | 2.52 |
| New Zealand | 430 | 1.92 |

World heat map:

```r
# Some modifications are needed
top_corresponding$country <- as.character(top_corresponding$country)
top_corresponding$country[top_corresponding$country == "United States"] <- "USA"
top_corresponding$country[top_corresponding$country == "United Kingdom"] <- "UK"
top_corresponding$country = as.factor(top_corresponding$country)

# Drawing the map
top_corrsponding_map = ggplot(top_corresponding) +
  geom_map(
    dat = world_map, map = world_map, aes(map_id = region),
    fill = "white", color = "#7f7f7f", size = 0.25
  ) +
  geom_map(map = world_map, aes(map_id = country, fill = log10(count)), size = 0.25) +
  scale_fill_gradient(low = "snow2", high = "red2", name = "Total corresponding authors (log
  expand_limits(x = world_map$long, y = world_map$lat) + theme(legend.position="bottom",
        axis.line=element_blank(),
        axis.text=element_blank(),
        axis.ticks=element_blank(),
        axis.title=element_blank(),
        panel.background=element_blank(),
        panel.border=element_blank(),
        panel.grid=element_blank())
```

```
top_corrsponding_map
```

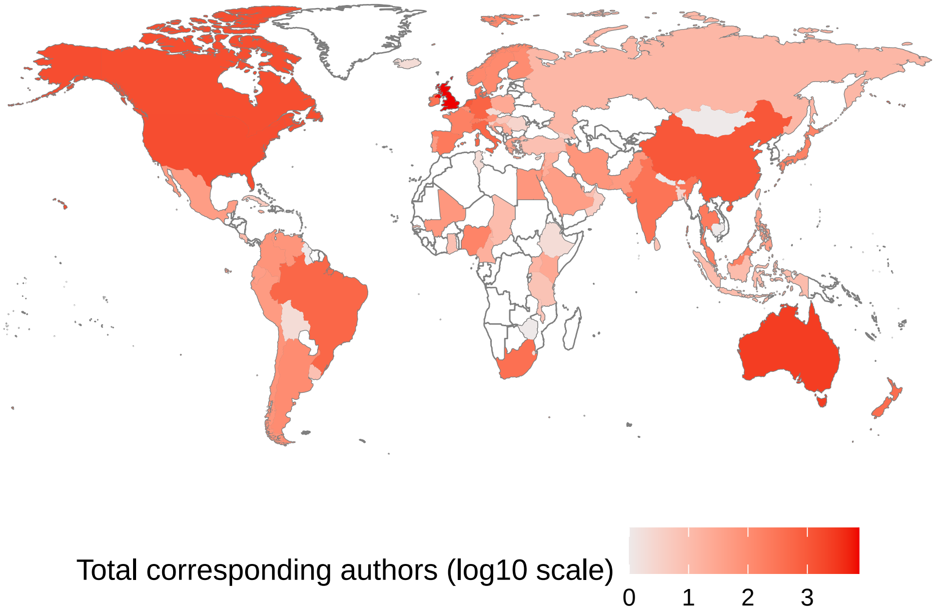

```
# ggsave("figures/Fig2B.png", width = 10, height = 5, dpi = 300)
```

###### 2.2.2.1 World Bank Income Regions

```
cochrane = cochrane %>% mutate(corresponding_author_region_income = ifelse(is.na(correspondi

kable(cochrane %>%
  filter(!is.na(corresponding_author_region_income)) %>%
  group_by(corresponding_author_region_income) %>%
  summarize(count = n()) %>%
  mutate(percentage = round(count / sum(count) * 100, 2)) %>%
  arrange(desc(count)))
```

| corresponding_author_region_income | count | percentage |
|---|---|---|
| 1. High income: OECD | 18096 | 81.50 |
| 3. Upper middle income | 3124 | 14.07 |
| 4. Lower middle income | 703 | 3.17 |
| 5. Low income | 142 | 0.64 |
| 2. High income: nonOECD | 138 | 0.62 |

###### 2.2.2.2 English-speaking

```
cochrane = cochrane %>% mutate(corresponding_author_english = ifelse(is.na(corresponding_autl

kable(cochrane %>%
  filter(!is.na(corresponding_author_english)) %>%
  group_by(corresponding_author_english) %>%
  summarize(count = n()) %>%
  mutate(percentage = round(count / sum(count) * 100, 2)) %>%
  arrange(desc(count)))
```

| corresponding_author_english | count | percentage |
|---|---|---|
| Yes | 13871 | 62.09 |
| No | 8469 | 37.91 |

###### 2.2.2.3 Spanish-speaking

```
cochrane = cochrane %>% mutate(corresponding_author_spanish = ifelse(is.na(corresponding_autl

kable(cochrane %>%
  filter(!is.na(corresponding_author_spanish)) %>%
  group_by(corresponding_author_spanish) %>%
  summarize(count = n()) %>%
  mutate(percentage = round(count / sum(count) * 100, 2)) %>%
  arrange(desc(count)))
```

| corresponding_author_spanish | count | percentage |
|---|---|---|
| FALSE | 21597 | 96.67 |
| TRUE | 743 | 3.33 |

##### 2.2.3 Last authors

The overall number of countries:

```
cochrane$last_author_country[cochrane$last_author_country == ""] <- NA

length(unique(cochrane$last_author_country))
```

```
[1] 94
```

Countries with the highest number of last authors:

```
top_last = cochrane %>%
  filter(!is.na(last_author_country)) %>%
  group_by(last_author_country) %>%
  summarize(count = n()) %>%
  mutate(percentage = round(count / sum(count) * 100, 2)) %>%
  arrange(desc(count))

names(top_last)[1] = c("country")

kable(top_last[1:10,])
```

| country | count | percentage |
|---|---|---|
| United Kingdom | 7720 | 34.71 |
| Australia | 2640 | 11.87 |
| United States | 1626 | 7.31 |
| Canada | 1598 | 7.19 |
| Netherlands | 1062 | 4.78 |
| China | 956 | 4.30 |
| Germany | 640 | 2.88 |
| Brazil | 531 | 2.39 |
| Italy | 520 | 2.34 |
| Denmark | 419 | 1.88 |

World heat map:

```
# Some modifications are needed
top_last$country <- as.character(top_last$country)
top_last$country[top_last$country == "United States"] <- "USA"
top_last$country[top_last$country == "United Kingdom"] <- "UK"
top_last$country = as.factor(top_last$country)

# Drawing the map
top_last_map = ggplot(top_last) +
  geom_map(
    dat = world_map, map = world_map, aes(map_id = region),
    fill = "white", color = "#7f7f7f", size = 0.25
```

```
  ) +
  geom_map(map = world_map, aes(map_id = country, fill = log10(count)), size = 0.25) +
  scale_fill_gradient(low = "snow2", high = "red2", na.value = "black", name = "Total last au
  expand_limits(x = world_map$long, y = world_map$lat) + theme(legend.position="bottom",
        axis.line=element_blank(),
        axis.text=element_blank(),
        axis.ticks=element_blank(),
        axis.title=element_blank(),
        panel.background=element_blank(),
        panel.border=element_blank(),
        panel.grid=element_blank())

top_last_map
```

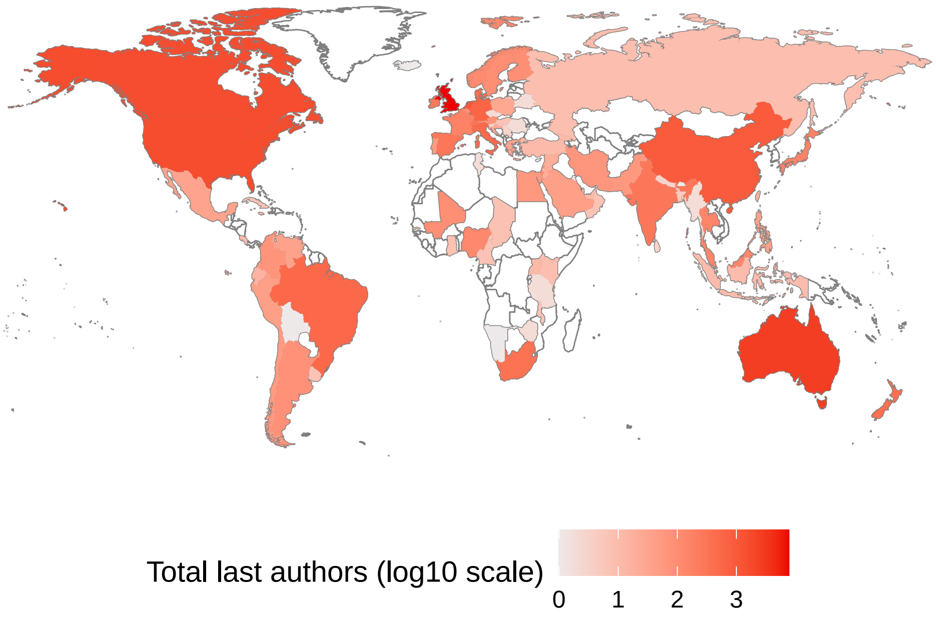

```
# ggsave("figures/Fig2C.png", width = 10, height = 5, dpi = 300)
```

###### 2.2.3.1 World Bank Income Regions

```
cochrane = cochrane %>% mutate(last_author_region_income = ifelse(is.na(last_author_country)

kable(cochrane %>%
  filter(!is.na(last_author_region_income)) %>%
  group_by(last_author_region_income) %>%
```

```
  summarize(count = n()) %>%
  mutate(percentage = round(count / sum(count) * 100, 2)) %>%
  arrange(desc(count)))
```

| last_author_region_income | count | percentage |
|---|---|---|
| 1. High income: OECD | 18556 | 83.90 |
| 3. Upper middle income | 2646 | 11.96 |
| 4. Lower middle income | 642 | 2.90 |
| 5. Low income | 146 | 0.66 |
| 2. High income: nonOECD | 126 | 0.57 |

###### 2.2.3.2 English-speaking

```
cochrane = cochrane %>% mutate(last_author_english = ifelse(is.na(last_author_country) == T,

kable(cochrane %>%
  filter(!is.na(last_author_english)) %>%
  group_by(last_author_english) %>%
  summarize(count = n()) %>%
  mutate(percentage = round(count / sum(count) * 100, 2)) %>%
  arrange(desc(count)))
```

| last_author_english | count | percentage |
|---|---|---|
| Yes | 14252 | 64.09 |
| No | 7987 | 35.91 |

###### 2.2.3.3 Spanish-speaking

```
cochrane = cochrane %>% mutate(last_author_spanish = ifelse(is.na(last_author_country) == T,

kable(cochrane %>%
  filter(!is.na(last_author_spanish)) %>%
  group_by(last_author_spanish) %>%
  summarize(count = n()) %>%
  mutate(percentage = round(count / sum(count) * 100, 2)) %>%
  arrange(desc(count)))
```

| last_author_spanish | count | percentage |
|---|---|---|
| FALSE | 21588 | 97.07 |
| TRUE | 651 | 2.93 |

###### 2.2.3.3.1 Combining all maps in one

```
library(ggpubr)
ggarrange(top_first_map, top_corrsponding_map, top_last_map,
                    labels = c("A", "B", "C"),
                    ncol = 1)
```

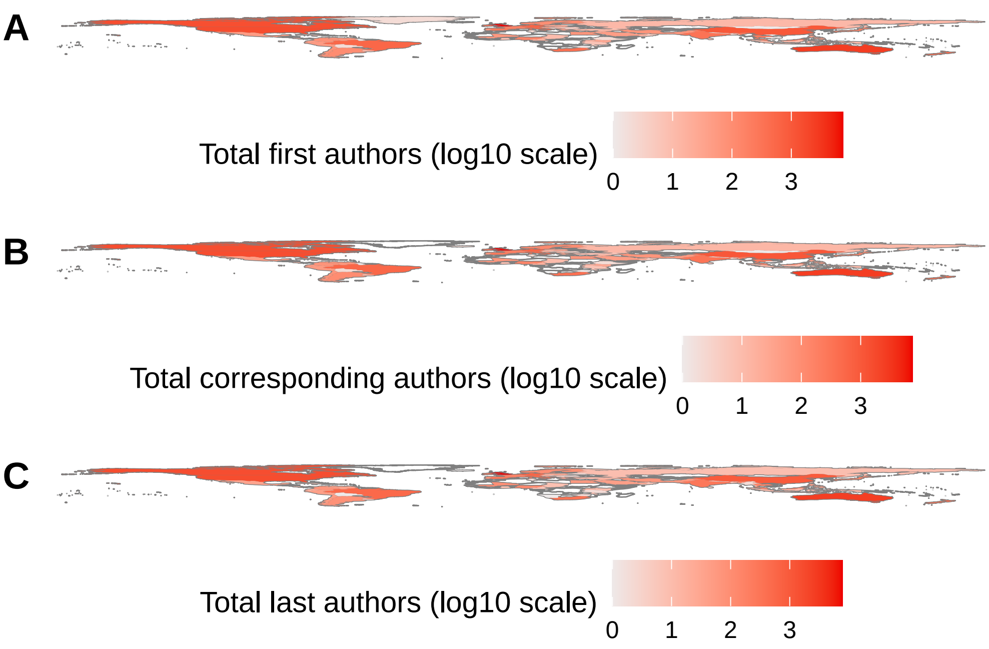

```
# ggsave("figures/Fig2.png", width = 10, height = 15, dpi = 300)
```

###### 2.2.3.3.2 Combining all information in one dataset

```
merged_first_corresponding = merge(top_first, top_corresponding, by = "country", all = T)
all_top = merge(merged_first_corresponding, top_last, by = "country", all = T)
all_top = all_top %>%
  mutate(across(where(is.numeric), ~coalesce(., 0)))
names(all_top) = c("country", "count_first", "percentage_first", "count_corrsponding", "perc

# write.csv(all_top, "figures/Appendix3.csv")
```

###### 2.2.3.3.3 Logistic regression

Income regions

```
log_first_income = glm(first_author_lowincome~year, data = cochrane, family = binomial(link="

log_cor_income = glm(corresponding_author_lowincome~year, data = cochrane, family = binomial

log_last_income = glm(last_author_lowincome~year, data = cochrane, family = binomial(link="lo

summary(log_first_income)
```

```
Call:
glm(formula = first_author_lowincome ~ year, family = binomial(link = "logit"),
    data = cochrane)

Coefficients:
              Estimate Std. Error z value Pr(>|z|)
(Intercept) -62.228165   5.884975  -10.57   <2e-16 ***
year          0.030205   0.002925   10.33   <2e-16 ***
---
Signif. codes:  0 '***' 0.001 '**' 0.01 '*' 0.05 '.' 0.1 ' ' 1

(Dispersion parameter for binomial family taken to be 1)

    Null deviance: 21430  on 22210  degrees of freedom
Residual deviance: 21322  on 22209  degrees of freedom
  (469 observations deleted due to missingness)
AIC: 21326

Number of Fisher Scoring iterations: 4
```

```
summary(log_cor_income)
```

```
Call:
glm(formula = corresponding_author_lowincome ~ year, family = binomial(link = "logit"),
    data = cochrane)

Coefficients:
              Estimate Std. Error z value Pr(>|z|)
```

```
(Intercept) -56.830314   5.875987  -9.672   <2e-16 ***
year          0.027514   0.002921   9.421   <2e-16 ***
---
Signif. codes:  0 '***' 0.001 '**' 0.01 '*' 0.05 '.' 0.1 ' ' 1

(Dispersion parameter for binomial family taken to be 1)

    Null deviance: 21264  on 22202  degrees of freedom
Residual deviance: 21174  on 22201  degrees of freedom
  (477 observations deleted due to missingness)
AIC: 21178

Number of Fisher Scoring iterations: 4
```

```
summary(log_last_income)
```

```
Call:
glm(formula = last_author_lowincome ~ year, family = binomial(link = "logit"),
    data = cochrane)

Coefficients:
              Estimate Std. Error z value Pr(>|z|)
(Intercept) -59.037059   6.287115  -9.390   <2e-16 ***
year          0.028526   0.003125   9.129   <2e-16 ***
---
Signif. codes:  0 '***' 0.001 '**' 0.01 '*' 0.05 '.' 0.1 ' ' 1

(Dispersion parameter for binomial family taken to be 1)

    Null deviance: 19518  on 22115  degrees of freedom
Residual deviance: 19434  on 22114  degrees of freedom
  (564 observations deleted due to missingness)
AIC: 19438

Number of Fisher Scoring iterations: 4
```

English-speaking

```
log_first_english = glm(relevel(factor(first_author_english), ref="Yes")~year, data = cochra
```

```r
log_cor_english = glm(relevel(factor(corresponding_author_english), ref="Yes")~year, data = c

log_last_english = glm(relevel(factor(last_author_english), ref="Yes")~year, data = cochrane

summary(log_first_english)
```

```
Call:
glm(formula = relevel(factor(first_author_english), ref = "Yes") ~
    year, family = binomial(link = "logit"), data = cochrane)

Coefficients:
              Estimate Std. Error z value Pr(>|z|)
(Intercept) -57.460953   4.692821  -12.24   <2e-16 ***
year          0.028324   0.002333   12.14   <2e-16 ***
---
Signif. codes:  0 '***' 0.001 '**' 0.01 '*' 0.05 '.' 0.1 ' ' 1

(Dispersion parameter for binomial family taken to be 1)

    Null deviance: 29700  on 22362  degrees of freedom
Residual deviance: 29551  on 22361  degrees of freedom
  (317 observations deleted due to missingness)
AIC: 29555

Number of Fisher Scoring iterations: 4
```

```r
summary(log_cor_english)
```

```
Call:
glm(formula = relevel(factor(corresponding_author_english), ref = "Yes") ~
    year, family = binomial(link = "logit"), data = cochrane)

Coefficients:
              Estimate Std. Error z value Pr(>|z|)
(Intercept) -52.655968   4.667754  -11.28   <2e-16 ***
year          0.025934   0.002321   11.18   <2e-16 ***
---
Signif. codes:  0 '***' 0.001 '**' 0.01 '*' 0.05 '.' 0.1 ' ' 1
```

```
(Dispersion parameter for binomial family taken to be 1)

    Null deviance: 29651  on 22339  degrees of freedom
Residual deviance: 29525  on 22338  degrees of freedom
  (340 observations deleted due to missingness)
AIC: 29529

Number of Fisher Scoring iterations: 4
```

```
summary(log_last_english)
```

```
Call:
glm(formula = relevel(factor(last_author_english), ref = "Yes") ~
    year, family = binomial(link = "logit"), data = cochrane)

Coefficients:
              Estimate Std. Error z value Pr(>|z|)
(Intercept) -52.062030   4.779618  -10.89   <2e-16 ***
year          0.025594   0.002376   10.77   <2e-16 ***
---
Signif. codes:  0 '***' 0.001 '**' 0.01 '*' 0.05 '.' 0.1 ' ' 1

(Dispersion parameter for binomial family taken to be 1)

    Null deviance: 29041  on 22238  degrees of freedom
Residual deviance: 28924  on 22237  degrees of freedom
  (441 observations deleted due to missingness)
AIC: 28928

Number of Fisher Scoring iterations: 4
```

Spanish-speaking

```
log_first_spanish = glm(first_author_spanish~year, data = cochrane, family = binomial(link="l

log_cor_spanish = glm(corresponding_author_spanish~year, data = cochrane, family = binomial(l

log_last_spanish = glm(last_author_spanish~year, data = cochrane, family = binomial(link="log

summary(log_first_spanish)
```

```
Call:
glm(formula = first_author_spanish ~ year, family = binomial(link = "logit"),
    data = cochrane)

Coefficients:
              Estimate Std. Error z value Pr(>|z|)
(Intercept) -40.396359  12.611330  -3.203  0.00136 **
year          0.018413   0.006268   2.937  0.00331 **
---
Signif. codes:  0 '***' 0.001 '**' 0.01 '*' 0.05 '.' 0.1 ' ' 1

(Dispersion parameter for binomial family taken to be 1)

    Null deviance: 6587.3  on 22362  degrees of freedom
Residual deviance: 6578.7  on 22361  degrees of freedom
  (317 observations deleted due to missingness)
AIC: 6582.7

Number of Fisher Scoring iterations: 6
```

```r
summary(log_cor_spanish)
```

```
Call:
glm(formula = corresponding_author_spanish ~ year, family = binomial(link = "logit"),
    data = cochrane)

Coefficients:
              Estimate Std. Error z value Pr(>|z|)
(Intercept) -31.802450  12.599496  -2.524   0.0116 *
year          0.014135   0.006263   2.257   0.0240 *
---
Signif. codes:  0 '***' 0.001 '**' 0.01 '*' 0.05 '.' 0.1 ' ' 1

(Dispersion parameter for binomial family taken to be 1)

    Null deviance: 6518.5  on 22339  degrees of freedom
Residual deviance: 6513.4  on 22338  degrees of freedom
  (340 observations deleted due to missingness)
AIC: 6517.4
```

```
Number of Fisher Scoring iterations: 6
```

```r
summary(log_last_spanish)
```

```
Call:
glm(formula = last_author_spanish ~ year, family = binomial(link = "logit"),
    data = cochrane)

Coefficients:
             Estimate Std. Error z value Pr(>|z|)
(Intercept) -29.56766   13.56032  -2.180   0.0292 *
year          0.01296    0.00674   1.922   0.0545 .
---
Signif. codes:  0 '***' 0.001 '**' 0.01 '*' 0.05 '.' 0.1 ' ' 1

(Dispersion parameter for binomial family taken to be 1)

    Null deviance: 5880.2  on 22238  degrees of freedom
Residual deviance: 5876.5  on 22237  degrees of freedom
  (441 observations deleted due to missingness)
AIC: 5880.5

Number of Fisher Scoring iterations: 6
```

#### 2.3 Gender diversity

##### 2.3.1 Overall

```r
cochrane = cochrane %>% mutate(first_author_female = ifelse(first_author_gender == "F", T, F
                               corresponding_author_female = ifelse(corresponding_author_gend
                               last_author_female = ifelse(last_author_gender == "F", T, F))

kable(cochrane %>%
  filter(!is.na(first_author_female)) %>%
  group_by(first_author_female) %>%
  summarize(count = n()) %>%
  mutate(percentage = round(count / sum(count) * 100, 2)) %>%
  arrange(desc(count)))
```

| first_author_female | count | percentage |
|---|---|---|
| TRUE | 10545 | 50.81 |
| FALSE | 10207 | 49.19 |

```
kable(cochrane %>%
  filter(!is.na(corresponding_author_female)) %>%
  group_by(corresponding_author_female) %>%
  summarize(count = n()) %>%
  mutate(percentage = round(count / sum(count) * 100, 2)) %>%
  arrange(desc(count)))
```

| corresponding_author_female | count | percentage |
|---|---|---|
| FALSE | 10720 | 51.87 |
| TRUE | 9947 | 48.13 |

```
kable(cochrane %>%
  filter(!is.na(last_author_female)) %>%
  group_by(last_author_female) %>%
  summarize(count = n()) %>%
  mutate(percentage = round(count / sum(count) * 100, 2)) %>%
  arrange(desc(count)))
```

| last_author_female | count | percentage |
|---|---|---|
| FALSE | 13106 | 62.68 |
| TRUE | 7804 | 37.32 |

##### 2.3.2 Trend

```
yearly_trend_gender =
        cochrane %>%
        select(year,
               first_author_female,
               corresponding_author_female,
               last_author_female) %>%
        gather("indicator", "value", -year) %>%
        count(year, indicator, value) %>%
```

```
        mutate(indicator = recode(indicator,
                                  first_author_female = "Female first authors",
                                  corresponding_author_female = "Female corresponding author
                                  last_author_female = "Female last authors")) %>%
        complete(indicator, value, year, fill = list(n = 0)) %>%
        group_by(year, indicator) %>%
        mutate(p = n / sum(n)) %>%
        filter(value) %>%
        ungroup()

plot_trend_gender =
        yearly_trend_gender %>%
        ggplot() +
        aes(x = year,
            y = p,
            group = indicator,
            color = indicator) +
        geom_line(size = 0.75) +
        scale_y_continuous(limits = c(0, 1),
                           labels = scales::percent) +
        scale_color_discrete(name = NULL) +
        scale_fill_discrete(breaks = c("Female first authors",
                                       "Female corresponding authors",
                                       "Female last authors")) +
        labs(   title="Gender diversity in Cochrane Reviews' authorship",
                y = "Proportion of female authors (%)",
                x = "Year") +
        theme(panel.grid.minor = element_blank(),
              legend.position = c(0.2, 0.8),
              axis.text.x = element_text(angle = 45, vjust = 0.9, hjust=1)
              )
#png("figures/plot_trend_region_english.png", width = 10, height = 7, units = "in", res = 80
plot_trend_gender
```

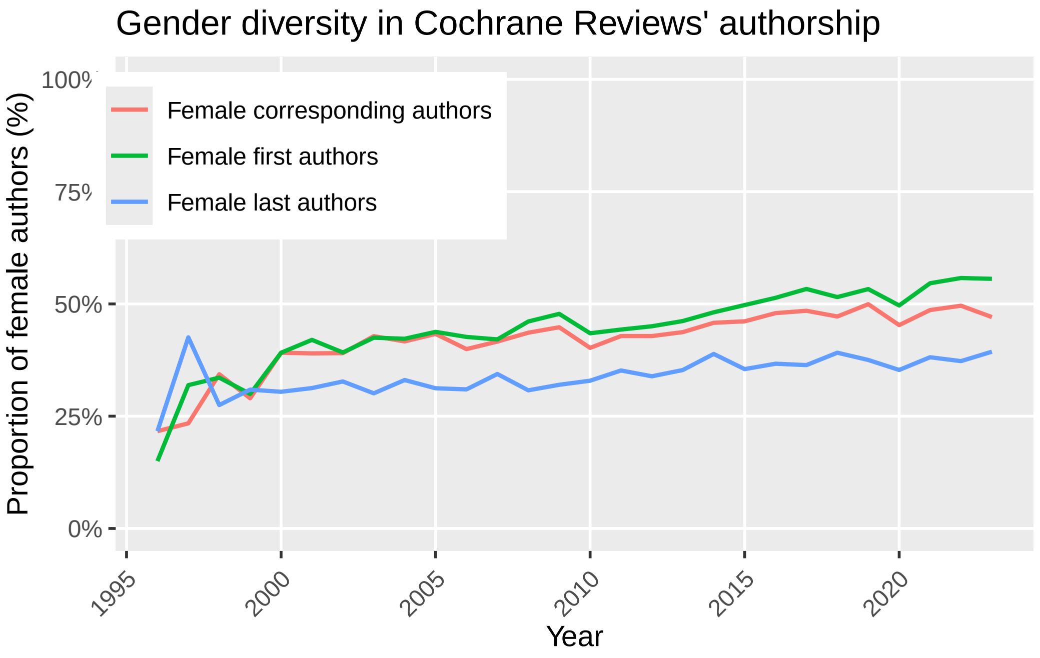

```
# ggsave("figures/Fig5.png", width = 10, height = 7, dpi = 300)
#dev.off()
```

###### 2.3.2.0.1 Logistic regression

```
log_first_female = glm(relevel(factor(first_author_gender), ref = "M")~year, data = cochrane

log_cor_female = glm(relevel(factor(corresponding_author_gender), ref = "M")~year, data = co

log_last_female = glm(relevel(factor(last_author_gender), ref = "M")~year, data = cochrane, 

summary(log_first_female)
```

```
Call:
glm(formula = relevel(factor(first_author_gender), ref = "M") ~
    year, family = binomial(link = "logit"), data = cochrane)

Coefficients:
              Estimate Std. Error z value Pr(>|z|)
(Intercept) -60.386139   4.667998  -12.94   <2e-16 ***
year          0.030040   0.002321   12.94   <2e-16 ***
```

```
---
Signif. codes:  0 '***' 0.001 '**' 0.01 '*' 0.05 '.' 0.1 ' ' 1

(Dispersion parameter for binomial family taken to be 1)

    Null deviance: 28763  on 20751  degrees of freedom
Residual deviance: 28594  on 20750  degrees of freedom
  (1928 observations deleted due to missingness)
AIC: 28598

Number of Fisher Scoring iterations: 3
```

```r
summary(log_cor_female)
```

```
Call:
glm(formula = relevel(factor(corresponding_author_gender), ref = "M") ~
    year, family = binomial(link = "logit"), data = cochrane)

Coefficients:
             Estimate Std. Error z value Pr(>|z|)
(Intercept) -43.07909    4.64674  -9.271   <2e-16 ***
year          0.02138    0.00231   9.255   <2e-16 ***
---
Signif. codes:  0 '***' 0.001 '**' 0.01 '*' 0.05 '.' 0.1 ' ' 1

(Dispersion parameter for binomial family taken to be 1)

    Null deviance: 28622  on 20666  degrees of freedom
Residual deviance: 28536  on 20665  degrees of freedom
  (2013 observations deleted due to missingness)
AIC: 28540

Number of Fisher Scoring iterations: 3
```

```r
summary(log_last_female)
```

```
Call:
glm(formula = relevel(factor(last_author_gender), ref = "M") ~
    year, family = binomial(link = "logit"), data = cochrane)
```

```
Coefficients:
              Estimate Std. Error z value Pr(>|z|)
(Intercept) -30.348906   4.813108  -6.305 2.87e-10 ***
year          0.014831   0.002393   6.198 5.72e-10 ***
---
Signif. codes:  0 '***' 0.001 '**' 0.01 '*' 0.05 '.' 0.1 ' ' 1

(Dispersion parameter for binomial family taken to be 1)

    Null deviance: 27628  on 20909  degrees of freedom
Residual deviance: 27590  on 20908  degrees of freedom
  (1770 observations deleted due to missingness)
AIC: 27594

Number of Fisher Scoring iterations: 4
```

##### 2.4 Diversity among review groups

Since some reviews were a collaboration between two review groups, we add a new column and only include those reviews which were from just one review group:

```
cochrane = cochrane %>%
  mutate(group_one = ifelse(str_count(group, "Cochrane") == 1, group, NA))
```

Then,

```
cochrane = cochrane %>% mutate(first_author_english_F = ifelse(first_author_english == "Yes"
                               corresponding_author_english_F = ifelse(corresponding_author_e
                               last_author_english_F = ifelse(last_author_english == "Yes", 

summary_table = cochrane %>%
    filter(is.na(group_one) == F) %>%
    group_by(group_one) %>%
    summarise(
        first_highincome = mean(first_author_lowincome, na.rm = TRUE),
        corresponding_highincome = mean(corresponding_author_lowincome, na.rm = TRUE),
        last_highincome = mean(last_author_lowincome, na.rm = TRUE),
        first_english = mean(first_author_english_F, na.rm = TRUE),
        corresponding_english = mean(corresponding_author_english_F, na.rm = TRUE),
```

```
        last_english = mean(last_author_english_F, na.rm = TRUE),
        first_spanish = mean(first_author_spanish, na.rm = TRUE),
        corresponding_spanish = mean(corresponding_author_spanish, na.rm = TRUE),
        last_spanish = mean(last_author_spanish, na.rm = TRUE),
        first_female = mean(first_author_female, na.rm = TRUE),
        corresponding_female = mean(corresponding_author_female, na.rm = TRUE),
        last_female = mean(last_author_female, na.rm = TRUE)
    ) %>%
    gather(key, value, -group_one) %>%
    separate(key, into = c("author", "indicator"), sep = "_")

# write.csv(summary_table, "figures/Appendix6.csv")
```

And now, plotting it:

```
plot_groups_diversity = summary_table %>%
        mutate(group_one = str_replace_all(group_one, c("Cochrane " = "", " Group" = ""))) %>
        ggplot() +
        aes(x = indicator,
            y = value,
            fill = author) +
        geom_bar(position = position_dodge(width = 0.7), width = 0.6, stat="identity") +
        facet_wrap(~ group_one, ncol = 10) +
        scale_y_continuous(limits = c(0, 1),
                           labels = scales::percent) +
        scale_color_discrete(name = NULL)  +
        labs(y = "Proportion of reviews\n",
             x = "") +
        theme(panel.grid.minor = element_blank(),
              legend.position = c(0.8, 0.01),
              axis.text.x = element_text(angle = 70, vjust = 0.9, hjust=1, size = 7))

plot_groups_diversity
```

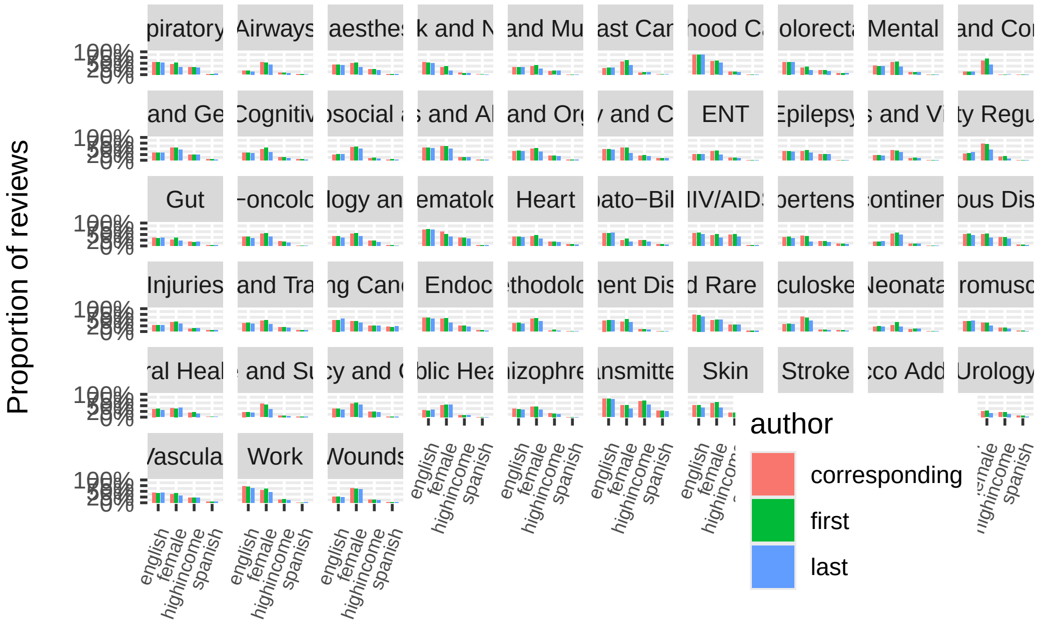

```
# ggsave("figures/Appendix5.png", width = 20, height = 12, dpi = 300)
```

Fisher exact test for first author:

```
fisher.test(table(cochrane$group_one, cochrane$first_author_lowincome), simulate.p.value = T
```

```
	Fisher's Exact Test for Count Data with simulated p-value (based on
	2000 replicates)

data:  table(cochrane$group_one, cochrane$first_author_lowincome)
p-value = 0.0004998
alternative hypothesis: two.sided
```

```
fisher.test(table(cochrane$group_one, cochrane$first_author_english), simulate.p.value = T)
```

```
	Fisher's Exact Test for Count Data with simulated p-value (based on
	2000 replicates)

data:  table(cochrane$group_one, cochrane$first_author_english)
p-value = 0.0004998
alternative hypothesis: two.sided
```

```r
fisher.test(table(cochrane$group_one, cochrane$first_author_female), simulate.p.value = T)
```

```
	Fisher's Exact Test for Count Data with simulated p-value (based on
	2000 replicates)

data:  table(cochrane$group_one, cochrane$first_author_female)
p-value = 0.0004998
alternative hypothesis: two.sided
```

Fisher exact test for corresponding author:

```r
fisher.test(table(cochrane$group_one, cochrane$corresponding_author_lowincome), simulate.p.va
```

```
	Fisher's Exact Test for Count Data with simulated p-value (based on
	2000 replicates)

data:  table(cochrane$group_one, cochrane$corresponding_author_lowincome)
p-value = 0.0004998
alternative hypothesis: two.sided
```

```r
fisher.test(table(cochrane$group_one, cochrane$corresponding_author_english), simulate.p.valu
```

```
	Fisher's Exact Test for Count Data with simulated p-value (based on
	2000 replicates)

data:  table(cochrane$group_one, cochrane$corresponding_author_english)
p-value = 0.0004998
alternative hypothesis: two.sided
```

```r
fisher.test(table(cochrane$group_one, cochrane$corresponding_author_female), simulate.p.value
```

```
	Fisher's Exact Test for Count Data with simulated p-value (based on
	2000 replicates)

data:  table(cochrane$group_one, cochrane$corresponding_author_female)
p-value = 0.0004998
alternative hypothesis: two.sided
```

Fisher exact test for last author:

```r
fisher.test(table(cochrane$group_one, cochrane$last_author_lowincome), simulate.p.value = T)
```

```

	Fisher's Exact Test for Count Data with simulated p-value (based on
	2000 replicates)

data:  table(cochrane$group_one, cochrane$last_author_lowincome)
p-value = 0.0004998
alternative hypothesis: two.sided
```

```r
fisher.test(table(cochrane$group_one, cochrane$last_author_english), simulate.p.value = T)
```

```

	Fisher's Exact Test for Count Data with simulated p-value (based on
	2000 replicates)

data:  table(cochrane$group_one, cochrane$last_author_english)
p-value = 0.0004998
alternative hypothesis: two.sided
```

```r
fisher.test(table(cochrane$group_one, cochrane$last_author_female), simulate.p.value = T)
```

```

	Fisher's Exact Test for Count Data with simulated p-value (based on
	2000 replicates)

data:  table(cochrane$group_one, cochrane$last_author_female)
p-value = 0.0004998
alternative hypothesis: two.sided
```

##### 2.4.1 Mixed-effects logistic regression

###### 2.4.1.1 First authorship

Gender:

```r
female_first_glmer = glmer(first_author_female ~ year + (1|group_one), data = cochrane, famil

summary(female_first_glmer)
```

```
Generalized linear mixed model fit by maximum likelihood (Adaptive
  Gauss-Hermite Quadrature, nAGQ = 10) [glmerMod]
 Family: binomial  ( logit )
Formula: first_author_female ~ year + (1 | group_one)
   Data: cochrane
Control: glmerControl(optimizer = "bobyqa", optCtrl = list(maxfun = 2e+05))

     AIC      BIC   logLik deviance df.resid
 27997.5  28021.3 -13995.7  27991.5    20726

Scaled residuals:
    Min      1Q  Median      3Q     Max
-1.8646 -0.9712  0.6447  0.9362  1.8956

Random effects:
 Groups    Name        Variance Std.Dev.
 group_one (Intercept) 0.1507   0.3882
Number of obs: 20729, groups:  group_one, 53

Fixed effects:
              Estimate Std. Error z value Pr(>|z|)
(Intercept) -67.290134   4.722042  -14.25   <2e-16 ***
year          0.033478   0.002347   14.26   <2e-16 ***
---
Signif. codes:  0 '***' 0.001 '**' 0.01 '*' 0.05 '.' 0.1 ' ' 1

Correlation of Fixed Effects:
     (Intr)
year -1.000
optimizer (bobyqa) convergence code: 0 (OK)
Model failed to converge with max|grad| = 0.104649 (tol = 0.002, component 1)
Model is nearly unidentifiable: very large eigenvalue
 - Rescale variables?
Model is nearly unidentifiable: large eigenvalue ratio
 - Rescale variables?
```

Region (income):

```r
low_first_glmer = glmer(first_author_lowincome ~ year + (1|group_one), data = cochrane, famil

summary(low_first_glmer)
```

```
Generalized linear mixed model fit by maximum likelihood (Adaptive
  Gauss-Hermite Quadrature, nAGQ = 10) [glmerMod]
 Family: binomial  ( logit )
Formula: first_author_lowincome ~ year + (1 | group_one)
   Data: cochrane
Control: glmerControl(optimizer = "bobyqa", optCtrl = list(maxfun = 2e+05))

     AIC      BIC   logLik deviance df.resid
 20425.2  20449.2 -10209.6  20419.2    22185

Scaled residuals:
    Min      1Q  Median      3Q     Max
-1.5414 -0.5237 -0.4101 -0.2847  5.2361

Random effects:
 Groups    Name        Variance Std.Dev.
 group_one (Intercept) 0.5306   0.7284
Number of obs: 22188, groups:  group_one, 53

Fixed effects:
              Estimate Std. Error z value Pr(>|z|)
(Intercept) -67.754217   6.146790  -11.02   <2e-16 ***
year          0.032895   0.003054   10.77   <2e-16 ***
---
Signif. codes:  0 '***' 0.001 '**' 0.01 '*' 0.05 '.' 0.1 ' ' 1

Correlation of Fixed Effects:
     (Intr)
year -1.000
optimizer (bobyqa) convergence code: 0 (OK)
Model failed to converge with max|grad| = 0.118185 (tol = 0.002, component 1)
Model is nearly unidentifiable: very large eigenvalue
 - Rescale variables?
Model is nearly unidentifiable: large eigenvalue ratio
 - Rescale variables?
```

Region (English):

```r
english_first_glmer = glmer(first_author_english_F ~ year + (1|group_one), data = cochrane, f

summary(english_first_glmer)
```

```
Generalized linear mixed model fit by maximum likelihood (Adaptive
  Gauss-Hermite Quadrature, nAGQ = 10) [glmerMod]
 Family: binomial  ( logit )
Formula: first_author_english_F ~ year + (1 | group_one)
   Data: cochrane
Control: glmerControl(optimizer = "bobyqa", optCtrl = list(maxfun = 2e+05))

     AIC      BIC   logLik deviance df.resid
 28109.7  28133.8 -14051.9  28103.7    22337

Scaled residuals:
    Min      1Q  Median      3Q     Max
-2.5494 -0.7827 -0.5523  1.0671  2.8028

Random effects:
 Groups    Name        Variance Std.Dev.
 group_one (Intercept) 0.4976   0.7054
Number of obs: 22340, groups:  group_one, 53

Fixed effects:
              Estimate Std. Error z value Pr(>|z|)
(Intercept) -62.631033   5.008177  -12.51   <2e-16 ***
year          0.030954   0.002489   12.44   <2e-16 ***
---
Signif. codes:  0 '***' 0.001 '**' 0.01 '*' 0.05 '.' 0.1 ' ' 1

Correlation of Fixed Effects:
     (Intr)
year -1.000
optimizer (bobyqa) convergence code: 0 (OK)
Model failed to converge with max|grad| = 0.0110084 (tol = 0.002, component 1)
Model is nearly unidentifiable: very large eigenvalue
 - Rescale variables?
Model is nearly unidentifiable: large eigenvalue ratio
 - Rescale variables?
```

Region (Spanish)

```r
spanish_first_glmer = glmer(first_author_spanish ~ year + (1|group_one), data = cochrane, fam

summary(spanish_first_glmer)
```

```
Generalized linear mixed model fit by maximum likelihood (Adaptive
  Gauss-Hermite Quadrature, nAGQ = 10) [glmerMod]
 Family: binomial  ( logit )
Formula: first_author_spanish ~ year + (1 | group_one)
   Data: cochrane
Control: glmerControl(optimizer = "bobyqa", optCtrl = list(maxfun = 2e+05))

     AIC      BIC   logLik deviance df.resid
  6339.2   6363.2  -3166.6   6333.2    22337

Scaled residuals:
    Min      1Q  Median      3Q     Max
-0.6005 -0.2171 -0.1674 -0.1231 11.2114

Random effects:
 Groups    Name        Variance Std.Dev.
 group_one (Intercept) 0.9084   0.9531
Number of obs: 22340, groups:  group_one, 53

Fixed effects:
              Estimate Std. Error z value Pr(>|z|)
(Intercept) -32.363947  10.255998  -3.156   0.0016 **
year          0.014307   0.005097   2.807   0.0050 **
---
Signif. codes:  0 '***' 0.001 '**' 0.01 '*' 0.05 '.' 0.1 ' ' 1

Correlation of Fixed Effects:
     (Intr)
year -1.000
optimizer (bobyqa) convergence code: 0 (OK)
Model failed to converge with max|grad| = 0.155527 (tol = 0.002, component 1)
Model is nearly unidentifiable: very large eigenvalue
 - Rescale variables?
Model is nearly unidentifiable: large eigenvalue ratio
 - Rescale variables?
```

###### 2.4.1.2 Corresponding authorship

Gender:

```r
female_corresponding_glmer = glmer(corresponding_author_female ~ year + (1|group_one), data =

summary(female_corresponding_glmer)
```

```
Generalized linear mixed model fit by maximum likelihood (Adaptive
  Gauss-Hermite Quadrature, nAGQ = 10) [glmerMod]
 Family: binomial  ( logit )
Formula: corresponding_author_female ~ year + (1 | group_one)
   Data: cochrane
Control: glmerControl(optimizer = "bobyqa", optCtrl = list(maxfun = 2e+05))

     AIC      BIC   logLik deviance df.resid
 27694.7  27718.5 -13844.4  27688.7    20642

Scaled residuals:
    Min      1Q  Median      3Q     Max
-1.7915 -0.9454 -0.5798  0.9416  1.8821

Random effects:
 Groups    Name        Variance Std.Dev.
 group_one (Intercept) 0.1867   0.4321
Number of obs: 20645, groups:  group_one, 53

Fixed effects:
              Estimate Std. Error z value Pr(>|z|)
(Intercept) -46.501394   4.776502  -9.735   <2e-16 ***
year          0.023094   0.002375   9.726   <2e-16 ***
---
Signif. codes:  0 '***' 0.001 '**' 0.01 '*' 0.05 '.' 0.1 ' ' 1

Correlation of Fixed Effects:
     (Intr)
year -1.000
optimizer (bobyqa) convergence code: 0 (OK)
Model failed to converge with max|grad| = 0.0088291 (tol = 0.002, component 1)
Model is nearly unidentifiable: very large eigenvalue
 - Rescale variables?
Model is nearly unidentifiable: large eigenvalue ratio
 - Rescale variables?
```

Region (income):

```r
low_corresponding_glmer = glmer(corresponding_author_lowincome ~ year + (1|group_one), data =

summary(low_corresponding_glmer)
```

```
Generalized linear mixed model fit by maximum likelihood (Adaptive
  Gauss-Hermite Quadrature, nAGQ = 10) [glmerMod]
 Family: binomial  ( logit )
Formula: corresponding_author_lowincome ~ year + (1 | group_one)
   Data: cochrane
Control: glmerControl(optimizer = "bobyqa", optCtrl = list(maxfun = 2e+05))

     AIC      BIC   logLik deviance df.resid
 20272.9  20296.9 -10133.5  20266.9    22177

Scaled residuals:
    Min      1Q  Median      3Q     Max
-1.4756 -0.5233 -0.4058 -0.2879  5.8727

Random effects:
 Groups    Name        Variance Std.Dev.
 group_one (Intercept) 0.566    0.7523
Number of obs: 22180, groups:  group_one, 53

Fixed effects:
              Estimate Std. Error z value Pr(>|z|)
(Intercept) -62.781230   6.080449  -10.32   <2e-16 ***
year          0.030405   0.003022   10.06   <2e-16 ***
---
Signif. codes:  0 '***' 0.001 '**' 0.01 '*' 0.05 '.' 0.1 ' ' 1

Correlation of Fixed Effects:
     (Intr)
year -1.000
optimizer (bobyqa) convergence code: 0 (OK)
Model failed to converge with max|grad| = 0.0200516 (tol = 0.002, component 1)
Model is nearly unidentifiable: very large eigenvalue
 - Rescale variables?
Model is nearly unidentifiable: large eigenvalue ratio
 - Rescale variables?
```

Region (language):

```r
english_corresponding_glmer = glmer(corresponding_author_english_F ~ year + (1|group_one), da

summary(english_corresponding_glmer)
```

```
Generalized linear mixed model fit by maximum likelihood (Adaptive
  Gauss-Hermite Quadrature, nAGQ = 10) [glmerMod]
 Family: binomial  ( logit )
Formula: corresponding_author_english_F ~ year + (1 | group_one)
   Data: cochrane
Control: glmerControl(optimizer = "bobyqa", optCtrl = list(maxfun = 2e+05))

     AIC      BIC   logLik deviance df.resid
 28111.8  28135.8 -14052.9  28105.8    22314

Scaled residuals:
    Min      1Q  Median      3Q     Max
-2.5471 -0.7853 -0.5551  1.0706  2.7844

Random effects:
 Groups    Name        Variance Std.Dev.
 group_one (Intercept) 0.4889   0.6992
Number of obs: 22317, groups:  group_one, 53

Fixed effects:
              Estimate Std. Error z value Pr(>|z|)
(Intercept) -57.915366   4.953394  -11.69   <2e-16 ***
year          0.028609   0.002462   11.62   <2e-16 ***
---
Signif. codes:  0 '***' 0.001 '**' 0.01 '*' 0.05 '.' 0.1 ' ' 1

Correlation of Fixed Effects:
     (Intr)
year -1.000
optimizer (bobyqa) convergence code: 0 (OK)
Model failed to converge with max|grad| = 0.0100902 (tol = 0.002, component 1)
Model is nearly unidentifiable: very large eigenvalue
 - Rescale variables?
Model is nearly unidentifiable: large eigenvalue ratio
 - Rescale variables?
```

Region (Spanish)

```r
spanish_corresponding_glmer = glmer(corresponding_author_spanish ~ year + (1|group_one), data

summary(spanish_corresponding_glmer)
```

```
Generalized linear mixed model fit by maximum likelihood (Adaptive
  Gauss-Hermite Quadrature, nAGQ = 10) [glmerMod]
 Family: binomial  ( logit )
Formula: corresponding_author_spanish ~ year + (1 | group_one)
   Data: cochrane
Control: glmerControl(optimizer = "bobyqa", optCtrl = list(maxfun = 2e+05))

     AIC      BIC   logLik deviance df.resid
  6256.0   6280.1  -3125.0   6250.0    22314

Scaled residuals:
    Min      1Q  Median      3Q     Max
-0.5963 -0.2155 -0.1669 -0.1241 11.4036

Random effects:
 Groups    Name        Variance Std.Dev.
 group_one (Intercept) 1.008    1.004
Number of obs: 22317, groups:  group_one, 53

Fixed effects:
              Estimate Std. Error z value Pr(>|z|)
(Intercept) -24.136537  10.913239  -2.212   0.0270 *
year          0.010195   0.005424   1.880   0.0602 .
---
Signif. codes:  0 '***' 0.001 '**' 0.01 '*' 0.05 '.' 0.1 ' ' 1

Correlation of Fixed Effects:
     (Intr)
year -1.000
optimizer (bobyqa) convergence code: 0 (OK)
Model failed to converge with max|grad| = 0.159043 (tol = 0.002, component 1)
Model is nearly unidentifiable: very large eigenvalue
 - Rescale variables?
Model is nearly unidentifiable: large eigenvalue ratio
 - Rescale variables?
```

###### 2.4.1.3 Last authorship

Gender:

```r
female_last_glmer = glmer(last_author_female ~ year + (1|group_one), data = cochrane, family

summary(female_last_glmer)
```

```
Generalized linear mixed model fit by maximum likelihood (Adaptive
  Gauss-Hermite Quadrature, nAGQ = 10) [glmerMod]
 Family: binomial  ( logit )
Formula: last_author_female ~ year + (1 | group_one)
   Data: cochrane
Control: glmerControl(optimizer = "bobyqa", optCtrl = list(maxfun = 2e+05))

     AIC      BIC   logLik deviance df.resid
 26735.4  26759.2 -13364.7  26729.4    20884

Scaled residuals:
    Min      1Q  Median      3Q     Max
-1.2924 -0.7746 -0.5907  1.1027  2.3146

Random effects:
 Groups    Name        Variance Std.Dev.
 group_one (Intercept) 0.2026   0.4501
Number of obs: 20887, groups:  group_one, 53

Fixed effects:
              Estimate Std. Error z value Pr(>|z|)
(Intercept) -33.741140   4.996663  -6.753 1.45e-11 ***
year          0.016500   0.002484   6.643 3.06e-11 ***
---
Signif. codes:  0 '***' 0.001 '**' 0.01 '*' 0.05 '.' 0.1 ' ' 1

Correlation of Fixed Effects:
     (Intr)
year -1.000
optimizer (bobyqa) convergence code: 0 (OK)
Model failed to converge with max|grad| = 0.0237944 (tol = 0.002, component 1)
Model is nearly unidentifiable: very large eigenvalue
 - Rescale variables?
Model is nearly unidentifiable: large eigenvalue ratio
 - Rescale variables?
```

Region (income):

```r
low_last_glmer = glmer(last_author_lowincome ~ year + (1|group_one), data = cochrane, family

summary(low_last_glmer)
```

```
Generalized linear mixed model fit by maximum likelihood (Adaptive
  Gauss-Hermite Quadrature, nAGQ = 10) [glmerMod]
 Family: binomial  ( logit )
Formula: last_author_lowincome ~ year + (1 | group_one)
   Data: cochrane
Control: glmerControl(optimizer = "bobyqa", optCtrl = list(maxfun = 2e+05))

     AIC      BIC   logLik deviance df.resid
 18689.0  18713.0  -9341.5  18683.0    22090

Scaled residuals:
    Min      1Q  Median      3Q     Max
-1.0824 -0.4777 -0.3827 -0.2719  6.0659

Random effects:
 Groups    Name        Variance Std.Dev.
 group_one (Intercept) 0.4444   0.6666
Number of obs: 22093, groups:  group_one, 53

Fixed effects:
              Estimate Std. Error z value Pr(>|z|)
(Intercept) -63.935702   6.551110   -9.76   <2e-16 ***
year          0.030894   0.003255    9.49   <2e-16 ***
---
Signif. codes:  0 '***' 0.001 '**' 0.01 '*' 0.05 '.' 0.1 ' ' 1

Correlation of Fixed Effects:
     (Intr)
year -1.000
optimizer (bobyqa) convergence code: 0 (OK)
Model failed to converge with max|grad| = 0.0447231 (tol = 0.002, component 1)
Model is nearly unidentifiable: very large eigenvalue
 - Rescale variables?
Model is nearly unidentifiable: large eigenvalue ratio
 - Rescale variables?
```

Region (language):

```r
english_last_glmer = glmer(last_author_english_F ~ year + (1|group_one), data = cochrane, fam

summary(english_last_glmer)
```

```
Generalized linear mixed model fit by maximum likelihood (Adaptive
  Gauss-Hermite Quadrature, nAGQ = 10) [glmerMod]
 Family: binomial  ( logit )
Formula: last_author_english_F ~ year + (1 | group_one)
   Data: cochrane
Control: glmerControl(optimizer = "bobyqa", optCtrl = list(maxfun = 2e+05))

     AIC      BIC   logLik deviance df.resid
 27528.9  27552.9 -13761.4  27522.9    22213

Scaled residuals:
    Min      1Q  Median      3Q     Max
-2.5173 -0.7450 -0.5543  1.0669  2.7675

Random effects:
 Groups    Name        Variance Std.Dev.
 group_one (Intercept) 0.466    0.6826
Number of obs: 22216, groups:  group_one, 53

Fixed effects:
              Estimate Std. Error z value Pr(>|z|)
(Intercept) -57.371213   4.997418  -11.48   <2e-16 ***
year          0.028296   0.002484   11.39   <2e-16 ***
---
Signif. codes:  0 '***' 0.001 '**' 0.01 '*' 0.05 '.' 0.1 ' ' 1

Correlation of Fixed Effects:
     (Intr)
year -1.000
optimizer (bobyqa) convergence code: 0 (OK)
Model failed to converge with max|grad| = 0.00979551 (tol = 0.002, component 1)
Model is nearly unidentifiable: very large eigenvalue
 - Rescale variables?
Model is nearly unidentifiable: large eigenvalue ratio
 - Rescale variables?
```

Region (Spanish)

```r
spanish_last_glmer = glmer(last_author_spanish ~ year + (1|group_one), data = cochrane, famil

summary(spanish_last_glmer)
```

```
Generalized linear mixed model fit by maximum likelihood (Adaptive
  Gauss-Hermite Quadrature, nAGQ = 10) [glmerMod]
 Family: binomial  ( logit )
Formula: last_author_spanish ~ year + (1 | group_one)
   Data: cochrane
Control: glmerControl(optimizer = "bobyqa", optCtrl = list(maxfun = 2e+05))

     AIC      BIC   logLik deviance df.resid
  5654.9   5678.9  -2824.4   5648.9    22213

Scaled residuals:
    Min      1Q  Median      3Q     Max
-0.5477 -0.1985 -0.1574 -0.1158 12.5613

Random effects:
 Groups    Name        Variance Std.Dev.
 group_one (Intercept) 0.9701   0.9849
Number of obs: 22216, groups:  group_one, 53

Fixed effects:
              Estimate Std. Error z value Pr(>|z|)
(Intercept) -24.494341  10.528197  -2.327   0.0200 *
year          0.010313   0.005232   1.971   0.0487 *
---
Signif. codes:  0 '***' 0.001 '**' 0.01 '*' 0.05 '.' 0.1 ' ' 1

Correlation of Fixed Effects:
     (Intr)
year -1.000
optimizer (bobyqa) convergence code: 0 (OK)
Model failed to converge with max|grad| = 0.13655 (tol = 0.002, component 1)
Model is nearly unidentifiable: very large eigenvalue
 - Rescale variables?
Model is nearly unidentifiable: large eigenvalue ratio
 - Rescale variables?
```

#### 2.5 Non-Cochrane reviews

First download using these:

```
# esearch -db pubmed -query '("Systematic Review"[PT]) NOT ("The Cochrane database of systema

# esearch -db pubmed -query '("Systematic Review"[PT]) NOT ("The Cochrane database of systema
```

New one:

```
# esearch -db pubmed -query '("Systematic Review"[PT]) NOT ("The Cochrane database of systema

# esearch -db pubmed -query '("Systematic Review"[PT]) NOT ("The Cochrane database of systema
```

Then:

```
pubmed_raw = readLines("data/pubmed/pubmed_aff.txt")

pmid = grep("^PMID- \\d+", pubmed_raw, value = TRUE)
pmid = sub("^PMID- ", "", pmid)

pmid_indices = grep("^PMID- \\d+", pubmed_raw)

dp = grep("^DP  -", pubmed_raw, value = TRUE)
dp = sub("^DP  - ", "", dp)

ad = gregexpr("^AD  - .+", pubmed_raw)
ad = regmatches(pubmed_raw, ad)
pmid_indices_plus2 = pmid_indices + 2
ad = ad[pmid_indices_plus2]
ad = sapply(ad, function(x) ifelse(length(x) == 0, NA, as.character(x)))
ad = sub("^AD  - ", "", ad)

pubmed_aff = data.frame(pmid = pmid, date = dp, aff1 = ad)
pubmed_aff = pubmed_aff[!duplicated(pubmed_aff$pmid), ]
```

PubMed authors:

```
pubmed_raw = readLines("data/pubmed/pubmed_author_names.txt")

pmid = grep("^PMID- \\d+", pubmed_raw, value = TRUE)
```

```
pmid = sub("^PMID- ", "", pmid)
pmid_indices = grep("^PMID- \\d+", pubmed_raw)

fau = gregexpr("^FAU - .+", pubmed_raw)
fau = regmatches(pubmed_raw, fau)
pmid_indices_plus1 = pmid_indices + 1
fau = fau[pmid_indices_plus1]
fau = sapply(fau, function(x) ifelse(length(x) == 0, NA, as.character(x)))
fau = sub("^FAU - ", "", fau)

pubmed_authors = data.frame(pmid = pmid, first_author = fau)
```

Combine these two:

```
pubmed = merge(pubmed_aff, pubmed_authors, by = "pmid")
```

Extracting year, country, regions, first name, and gender:

```
# Year:
pubmed$year = as.integer(substr(pubmed$date, 1, 4))

# Country:
library(maps)

world <- rnaturalearth::ne_countries(returnclass = "sf")
world <- world %>% st_drop_geometry()
world <- world %>% select(name, region_un, region_wb, income_grp)

all_countries = str_c(unique(world.cities$country.etc), collapse = "|")
all_countries = paste0(all_countries, "|United States|United Kingdom")

pubmed$country_aff1 =
  sapply(str_extract_all(pubmed$aff1, all_countries),
         toString)

georgias = c("Georgia, USA", "Georgia, Georgia, USA", "Georgia, United States", "Georgia, US

for (pattern in georgias) {
  pubmed$country_aff1[pubmed$country_aff1 == pattern] <- "USA"
  }
```

```r
pubmed$country_aff1 = sapply(strsplit(pubmed$country_aff1, ",\\s*"), function(x) ifelse(leng

# Income region

### Chnaging UK to United Kingdom and USA to United States
pubmed$country_aff1 = ifelse(pubmed$country_aff1 == "UK",
                                  "United Kingdom",
                                  pubmed$country_aff1)

pubmed$country_aff1 <- ifelse(pubmed$country_aff1 == "USA",
                                  "United States",
                                  pubmed$country_aff1)

for (i in 1:nrow(pubmed)) {
  a = pubmed$country_aff1[i]
  b = world[grep(a, world$name), 4]
  ifelse(rlang::is_empty(b) == TRUE,
         pubmed$region_income_aff1[i] <- NA,
         pubmed$region_income_aff1[i] <- b)
}

# English speaking
english_speaking <- "United States|US|USA|United Kingdom|UK|England|Canada|Australia|Ireland

pubmed$english_aff1 = sapply(
  pubmed$country_aff1,
  function(x) ifelse(str_extract_all(x, english_speaking) == "character(0)",
                     F,
                     T))

# Soanish speaking
spanish_speaking <- "Argentina|Bolivia|Chile|Colombia|Costa Rica|Cuba|Dominican Republic|Ecua

pubmed$spanish_aff1 = sapply(
  pubmed$country_aff1,
  function(x) ifelse(str_extract_all(x, spanish_speaking) == "character(0)",
                     F,
                     T))

# Gender
```

```
pubmed$first_author_given_name = sapply(
  strsplit(
    pubmed$first_author, ",\\s*"), function(x) {
    if (length(x) == 2) {
        # Use regular expression to capture the first word after the comma
        match_result <- regmatches(x[2], regexec("\\b\\w+\\b", x[2]))
        if (!is.na(match_result[[1]]) && nchar(match_result[[1]]) > 1) {
            return(match_result[[1]])
        } else {
            return(NA)
        }
    } else {
        return(NA)
    }
})

genderdb = read.csv("data/wgnd_2_0_name-gender-code.csv")
genderdb = genderdb[order(genderdb$wgt, decreasing = TRUE), ]
genderdb = genderdb[!duplicated(genderdb$name), ]
genderdb$wgt = NULL
genderdb$code = NULL

pubmed$first_author_given_name_lower = tolower(pubmed$first_author_given_name)

pubmed = merge(pubmed, genderdb, by.x = "first_author_given_name_lower", by.y = "name", all.

pubmed$gender[pubmed$gender == "?"] <- NA

pubmed$first_author_given_name_lower = NULL
```

Save it:

```
# write.csv(pubmed, "data/pubmed/pubmed_final.csv")
```

Trend:

```
pubmed = pubmed %>% mutate(first_author_female = ifelse(gender == "F", T, F),
                               first_author_lowincome = ifelse(region_income_aff1 == "1. Hig
                           first_author_non_english = ifelse(english_aff1 == T, F, T),
```

```
                           first_author_spanish = ifelse(spanish_aff1 == T, T, F))

yearly_trend_pubmed =
        pubmed %>%
        select(year,
               first_author_female,
               first_author_lowincome,
               first_author_non_english,
               first_author_spanish) %>%
        gather("indicator", "value", -year) %>%
        count(year, indicator, value) %>%
        mutate(indicator = recode(indicator,
                                  first_author_female = "Female first authors",
                                  first_author_lowincome = "First author from non-high-income
                                  first_author_non_english = "First author from non-English s
                                  first_author_spanish = "First author from Spanish speaking
        complete(indicator, value, year, fill = list(n = 0)) %>%
        group_by(year, indicator) %>%
        mutate(p = n / sum(n)) %>%
        filter(value) %>%
        ungroup()

plot_trend_pubmed =
        yearly_trend_pubmed %>%
        filter(year >= 1996, year <= 2023) %>%
        ggplot() +
        aes(x = year,
            y = p,
            group = indicator,
            color = indicator) +
        geom_line(size = 0.75) +
        scale_y_continuous(limits = c(0, 1),
                           labels = scales::percent) +
        scale_color_discrete(name = NULL) +
        scale_fill_discrete(breaks = c("Female first authors",
                                       "First author from non-high-income OECD countries",
                                       "First author from non-English speaking countries",
                                       "First author from Spanish speaking countries")) +
        labs(title="Geographical and gender diversity in non-Cochrane reviews' authorship",
                y = "Proportion (%)",
```

```
                x = "Year") +
        theme(panel.grid.minor = element_blank(),
              legend.position = c(0.2, 0.8),
              axis.text.x = element_text(angle = 45, vjust = 0.9, hjust=1)
              )

plot_trend_pubmed
```

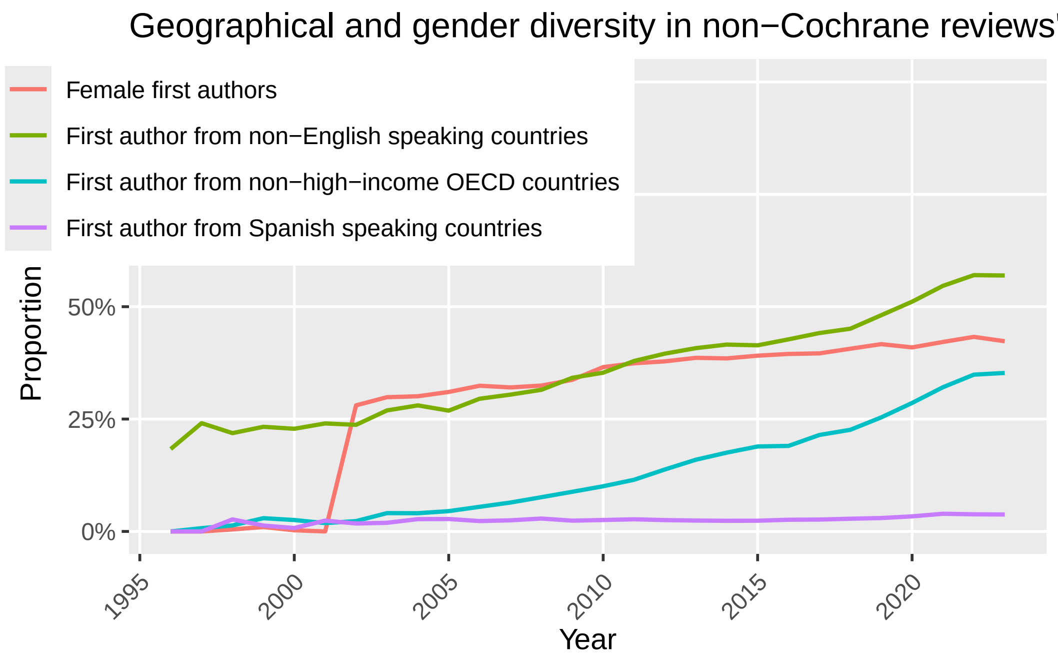

```
# ggsave("figures/Fig6.png", width = 10, height = 7, dpi = 300)
```

Testing two time series:

Non-high income:

Correlation:

```
dat = data.frame(cochrane = yearly_trend_region_lowincome %>% filter(indicator == "First autl

names(dat) = c("cochrane", "pubmed")

cor.test(dat$cochrane, dat$pubmed)
```

```
	Pearson's product-moment correlation

data:  dat$cochrane and dat$pubmed
t = 4.3348, df = 26, p-value = 0.0001944
alternative hypothesis: true correlation is not equal to 0
95 percent confidence interval:
 0.3621300 0.8221192
sample estimates:
     cor
0.647701
```

Granger Causality Test:

```
granger_test_result = grangertest(cochrane ~ pubmed, order = 3, data = dat)
summary(granger_test_result)
```

```
     Res.Df            Df           F               Pr(>F)
 Min.   :18.00   Min.   :-3   Min.   :0.7809   Min.   :0.52
 1st Qu.:18.75   1st Qu.:-3   1st Qu.:0.7809   1st Qu.:0.52
 Median :19.50   Median :-3   Median :0.7809   Median :0.52
 Mean   :19.50   Mean   :-3   Mean   :0.7809   Mean   :0.52
 3rd Qu.:20.25   3rd Qu.:-3   3rd Qu.:0.7809   3rd Qu.:0.52
 Max.   :21.00   Max.   :-3   Max.   :0.7809   Max.   :0.52
                 NA's   :1    NA's   :1        NA's   :1
```

non-English-speaking:

Correlation:

```
dat = data.frame(cochrane = yearly_trend_region_non_english %>% filter(indicator == "First au

names(dat) = c("cochrane", "pubmed")

cor.test(dat$cochrane, dat$pubmed)
```

```
	Pearson's product-moment correlation

data:  dat$cochrane and dat$pubmed
t = 7.3766, df = 26, p-value = 7.82e-08
alternative hypothesis: true correlation is not equal to 0
```

```
95 percent confidence interval:
 0.6485692 0.9149022
sample estimates:
      cor
0.8226038
```

Granger Causality Test:

```r
granger_test_result = grangertest(cochrane ~ pubmed, order = 3, data = dat)
summary(granger_test_result)
```

```
     Res.Df           Df           F                Pr(>F)
 Min.   :18.00   Min.   :-3   Min.   :0.4797   Min.   :0.7005
 1st Qu.:18.75   1st Qu.:-3   1st Qu.:0.4797   1st Qu.:0.7005
 Median :19.50   Median :-3   Median :0.4797   Median :0.7005
 Mean   :19.50   Mean   :-3   Mean   :0.4797   Mean   :0.7005
 3rd Qu.:20.25   3rd Qu.:-3   3rd Qu.:0.4797   3rd Qu.:0.7005
 Max.   :21.00   Max.   :-3   Max.   :0.4797   Max.   :0.7005
                 NA's   :1    NA's   :1        NA's   :1
```

Spanish-speaking:

Correlation:

```r
dat = data.frame(cochrane = yearly_trend_region_spanish %>% filter(indicator == "First autho

names(dat) = c("cochrane", "pubmed")

cor.test(dat$cochrane, dat$pubmed)
```

```
    Pearson's product-moment correlation

data:  dat$cochrane and dat$pubmed
t = 4.18, df = 26, p-value = 0.000292
alternative hypothesis: true correlation is not equal to 0
95 percent confidence interval:
 0.3417171 0.8144212
sample estimates:
      cor
0.6339711
```

Granger Causality Test:

```
granger_test_result = grangertest(cochrane ~ pubmed, order = 3, data = dat)
summary(granger_test_result)
```

```
     Res.Df            Df           F              Pr(>F)
 Min.   :18.00   Min.   :-3   Min.   :0.8526   Min.   :0.4833
 1st Qu.:18.75   1st Qu.:-3   1st Qu.:0.8526   1st Qu.:0.4833
 Median :19.50   Median :-3   Median :0.8526   Median :0.4833
 Mean   :19.50   Mean   :-3   Mean   :0.8526   Mean   :0.4833
 3rd Qu.:20.25   3rd Qu.:-3   3rd Qu.:0.8526   3rd Qu.:0.4833
 Max.   :21.00   Max.   :-3   Max.   :0.8526   Max.   :0.4833
                 NA's   :1    NA's   :1        NA's   :1
```

Gender:

Correlation:

```
dat = data.frame(cochrane = yearly_trend_gender %>% filter(indicator == "Female first authors

names(dat) = c("cochrane", "pubmed")

cor.test(dat$cochrane, dat$pubmed)
```

```
    Pearson's product-moment correlation

data:  dat$cochrane and dat$pubmed
t = 7.548, df = 26, p-value = 5.172e-08
alternative hypothesis: true correlation is not equal to 0
95 percent confidence interval:
 0.6594174 0.9179384
sample estimates:
      cor
0.8286382
```

Granger Causality Test:

```
granger_test_result = grangertest(cochrane ~ pubmed, order = 3, data = dat)
summary(granger_test_result)
```

```
    Res.Df            Df            F              Pr(>F)
 Min.   :18.00   Min.   :-3   Min.   :0.8207   Min.   :0.4993
 1st Qu.:18.75   1st Qu.:-3   1st Qu.:0.8207   1st Qu.:0.4993
 Median :19.50   Median :-3   Median :0.8207   Median :0.4993
 Mean   :19.50   Mean   :-3   Mean   :0.8207   Mean   :0.4993
 3rd Qu.:20.25   3rd Qu.:-3   3rd Qu.:0.8207   3rd Qu.:0.4993
 Max.   :21.00   Max.   :-3   Max.   :0.8207   Max.   :0.4993
                 NA's   :1    NA's   :1        NA's   :1
```
