## Appendix 3 for "Geographical and Gender Diversity in Cochrane and non-Cochrane Reviews Authorship: A Meta-Research Study"

**Appendix 3.** The list of the high-income, English-speaking, and Spanish-speaking countries.

| **High-income** | **English-speaking** | **Spanish-speaking** |
| --- | --- | --- |
| Antarctica | Australia | Argentina |
| Australia | Canada | Bolivia |
| Austria | Ireland | Chile |
| Bahamas | New Zealand | Colombia |
| Belgium | The United Kingdom | Costa Rica |
| Brunei | The United States | Cuba |
| Canada |  | Dominican Republic |
| Croatia |  | Ecuador |
| Cyprus |  | El Salvador |
| Czechia |  | Equatorial Guinea |
| Denmark |  | Guatemala |
| Estonia |  | Honduras |
| Eq. Guinea |  | Mexico |
| Falkland Is. |  | Nicaragua |
| Finland |  | Panama |
| France |  | Paraguay |
| Fr. S. Antarctic Lands |  | Peru |
| Germany |  | Puerto Rico |
| Greece |  | Spain |
| Greenland |  | Uruguay |
| Hungary |  | Venezuela |
| Iceland |  |  |
| Ireland |  |  |
| Israel |  |  |
| Italy |  |  |
| Japan |  |  |
| Kuwait |  |  |
| Luxembourg |  |  |
| The Netherlands |  |  |
| New Caledonia |  |  |
| New Zealand |  |  |
| Norway |  |  |
| Oman |  |  |
| Poland |  |  |
| Portugal |  |  |
| Puerto Rico |  |  |
| Qatar |  |  |
| Saudi Arabia |  |  |
| Slovakia |  |  |
| Slovenia |  |  |
| South Korea |  |  |
| Spain |  |  |
| Sweden |  |  |
| Switzerland |  |  |
| Taiwan |  |  |
| Trinidad and Tobago |  |  |
| United Arab Emirates |  |  |
| The United Kingdom |  |  |
| The United States |  |  |
