## Supplementary figures and images for "Geographical and Gender Diversity in Cochrane and non-Cochrane Reviews Authorship: A Meta-Research Study"

### Appendix 5

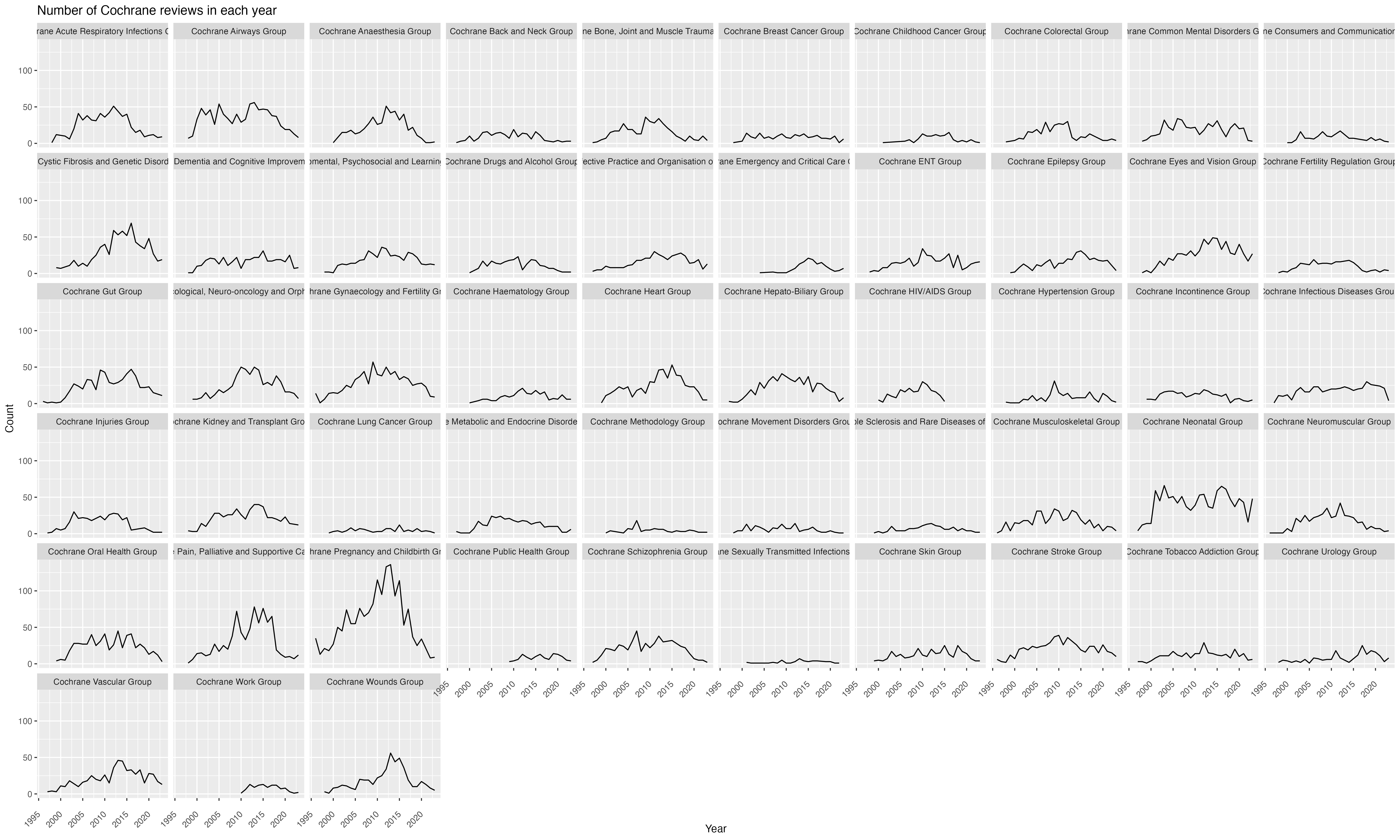

### Appendix 8

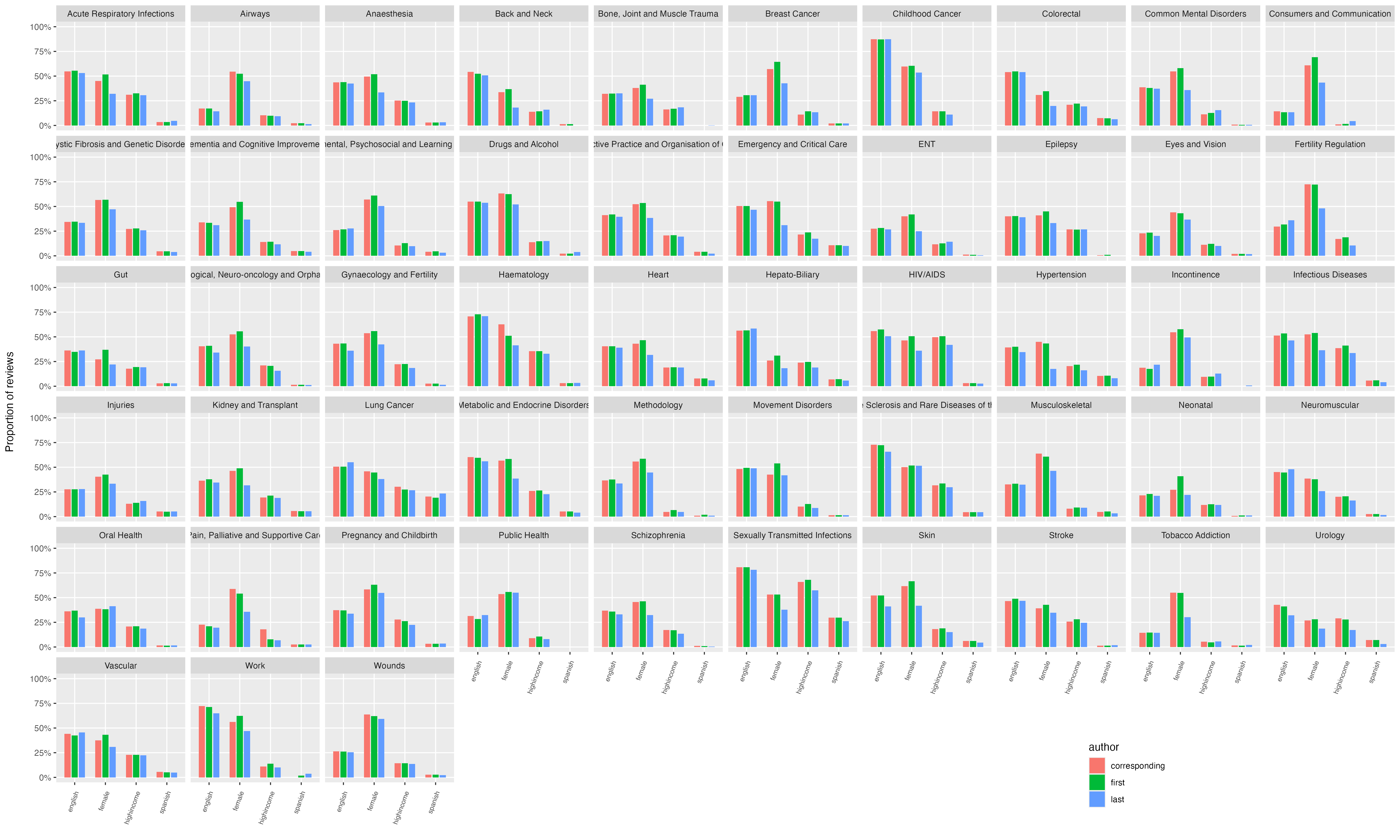
