## Appendix 7 for "Geographical and Gender Diversity in Cochrane and non-Cochrane Reviews Authorship: A Meta-Research Study"

**Appendix 6.** The outputs from the logistic regression models.

| glm(formula = first_author_lowincome ~ year, family = binomial(link = "logit"), data = cochrane)  AIC: 22542 | | | | |
| --- | --- | --- | --- | --- |
| **Variable** | **B** | **Exp(B)** | **SE** | **P** |
| year | 0.012 | 1.012 | 0.003 | <0.001 |
| Intercept | -25.386 | <0.001 | 5.591 | <0.001 |
| glm(formula = corresponding_author_lowincome ~ year, family = binomial(link = "logit"), data = cochrane)  AIC: 22428 | | | | |
| year | 0.013 | 1.013 | 0.003 | <0.001 |
| Intercept | -27.829 | <0.001 | 5.612 | <0.001 |
| glm(formula = last_author_lowincome ~ year, family = binomial(link = "logit"), data = cochrane)  AIC: 21127 | | | | |
| year | -0.003 | 0.997 | 0.003 | 0.262 |
| Intercept | 4.988 | 146.643 | 5.815 | 0.391 |
| glm(formula = relevel(factor(first_author_english), ref = "Yes") ~ year, family = binomial(link = "logit"), data = cochrane)  AIC: 29555 | | | | |
| year | 0.028 | 1.029 | 0.002 | <0.001 |
| Intercept | -57.461 | <0.001 | 4.693 | <0.001 |
| glm(formula = relevel(factor(corresponding_author_english), ref = "Yes") ~ year, family = binomial(link = "logit"), data = cochrane)  AIC: 29529 | | | | |
| year | 0.026 | 1.026 | 0.002 | <0.001 |
| Intercept | -52.656 | <0.001 | 4.668 | <0.001 |
| glm(formula = relevel(factor(last_author_english), ref = "Yes") ~ year, family = binomial(link = "logit"), data = cochrane)  AIC: 28928 | | | | |
| year | 0.026 | 1.026 | 0.002 | <0.001 |
| Intercept | -52.062 | <0.001 | 4.780 | <0.001 |
| glm(formula = first_author_spanish ~ year, family = binomial(link = "logit"), data = cochrane)  AIC: 6583 | | | | |
| year | 0.018 | 1.019 | 0.006 | 0.003 |
| Intercept | -40.396 | <0.001 | 12.611 | 0.001 |
| glm(formula = corresponding_author_spanish ~ year, family = binomial(link = "logit"), data = cochrane)  AIC: 6517 | | | | |
| year | 0.014 | 1.014 | 0.006263 | 0.024 |
| Intercept | -31.802 | <0.001 | 12.599 | 0.012 |
| glm(formula = last_author_spanish ~ year, family = binomial(link = "logit"), data = cochrane)  AIC: 5881 | | | | |
| year | 0.013 | 1.013 | 0.00674 | 0.055 |
| Intercept | -29.568 | <0.001 | 13.560 | 0.029 |
| glm(formula = relevel(factor(first_author_gender), ref = "M") ~ year, family = binomial(link = "logit"), data = cochrane)  AIC: 28598 | | | | |
| year | 0.030 | 1.030 | 0.002 | <0.001 |
| Intercept | -60.386 | <0.001 | 4.668 | <0.001 |
| glm(formula = relevel(factor(corresponding_author_gender), ref = "M") ~ year, family = binomial(link = "logit"), data = cochrane)  AIC: 28540 | | | | |
| year | 0.021 | 1.021 | 0.002 | <0.001 |
| Intercept | -43.079 | <0.001 | 4.647 | <0.001 |
| glm(formula = relevel(factor(last_author_gender), ref = "M") ~ year, family = binomial(link = "logit"), data = cochrane)  AIC: 27594 | | | | |
| year | 0.015 | 1.015 | 0.002 | <0.001 |
| Intercept | -30.349 | <0.001 | 4.813 | <0.001 |
