## Appendix 10 for "Geographical and Gender Diversity in Cochrane and non-Cochrane Reviews Authorship: A Meta-Research Study"

**Appendix 9.** The outputs from the random effect logistic regression models.

| glmer(first_author_lowincome ~ year + (1\|group_one), data = cochrane, family = binomial, control = glmerControl(optimizer = "bobyqa", optCtrl=list(maxfun=2e5)), nAGQ = 10)  AIC: 21698  Random effect variance (SD): 0.425 (0.6515) | | | | |
| --- | --- | --- | --- | --- |
| **Variable** | **B** | **Exp(B)** | **SE** | **P** |
| year | 0.013 | 1.013 | 0.003 | <0.001 |
| Intercept | -27.692 | <0.001 | 5.754 | <0.001 |
| glmer(corresponding_author_lowincome ~ year + (1\|group_one), data = cochrane, family = binomial, control = glmerControl(optimizer = "bobyqa", optCtrl=list(maxfun=2e5)), nAGQ = 10)  AIC: 21628  Random effect variance (SD): 0.441 (0.6641) | | | | |
| year | 0.014 | 1.014 | 0.003 | <0.001 |
| Intercept | -30.412 | <0.001 | 5.851 | <0.001 |
| glmer(last_author_lowincome ~ year + (1\|group_one), data = cochrane, family = binomial, control = glmerControl(optimizer = "bobyqa", optCtrl=list(maxfun=2e5)), nAGQ = 10)  AIC: 20494  Random effect variance (SD): 0.326 (0.5711) | | | | |
| year | -0.003 | 0.997 | 0.003 | 0.241 |
| Intercept | 4.998 | 148.117 | 5.648 | 0.376 |
| glmer(first_author_english_F ~ year + (1\|group_one), data = cochrane, family = binomial, control = glmerControl(optimizer = "bobyqa", optCtrl=list(maxfun=2e5)), nAGQ = 10)  AIC: 28110  Random effect variance (SD): 0.498 (0.7054) | | | | |
| year | 0.031 | 1.031 | 0.002 | <0.001 |
| Intercept | -62.631 | <0.001 | 5.008 | <0.001 |
| glmer(corresponding_author_english_F ~ year + (1\|group_one), data = cochrane, family = binomial, control = glmerControl(optimizer = "bobyqa", optCtrl=list(maxfun=2e5)), nAGQ = 10)  AIC: 28112  Random effect variance (SD): 0.489 (0.6992) | | | | |
| year | 0.029 | 1.029 | 0.002 | <0.001 |
| Intercept | -57.915 | <0.001 | 4.953 | <0.001 |
| glmer(last_author_english_F ~ year + (1\|group_one), data = cochrane, family = binomial, control = glmerControl(optimizer = "bobyqa", optCtrl=list(maxfun=2e5)), nAGQ = 10)  AIC: 27529  Random effect variance (SD): 0.466 (0.6826) | | | | |
| year | 0.028 | 1.028 | 0.002 | <0.001 |
| Intercept | -57.371 | <0.001 | 4.997 | <0.001 |
| glmer(first_author_spanish ~ year + (1\|group_one), data = cochrane, family = binomial, control = glmerControl(optimizer = "bobyqa", optCtrl=list(maxfun=2e5)), nAGQ = 10)  AIC: 6339  Random effect variance (SD): 0.908 (0.9531) | | | | |
| year | 0.014 | 1.014 | 0.005 | 0.005 |
| Intercept | -32.364 | <0.001 | 10.256 | 0.002 |
| glmer(corresponding_author_spanish ~ year + (1\|group_one), data = cochrane, family = binomial, control = glmerControl(optimizer = "bobyqa", optCtrl=list(maxfun=2e5)), nAGQ = 10)  AIC: 6256  Random effect variance (SD): 1.008 (1.004) | | | | |
| year | 0.010 | 1.010 | 0.005 | 0.060 |
| Intercept | -24.137 | <0.001 | 10.913 | 0.027 |
| glmer(last_author_spanish ~ year + (1\|group_one), data = cochrane, family = binomial, control = glmerControl(optimizer = "bobyqa", optCtrl=list(maxfun=2e5)), nAGQ = 10)  AIC: 5655  Random effect variance (SD): 0.970 (0.9849) | | | | |
| year | 0.010 | 1.010 | 0.005 | 0.049 |
| Intercept | -24.494 | <0.001 | 10.528 | 0.020 |
| glmer(first_author_female ~ year + (1\|group_one), data = cochrane, family = binomial, control = glmerControl(optimizer = "bobyqa", optCtrl=list(maxfun=2e5)), nAGQ = 10)  AIC: 27998  Random effect variance (SD): 0.151 (0.3882) | | | | |
| year | 0.033 | 1.033 | 0.002 | <0.001 |
| Intercept | -67.290 | <0.001 | 4.722 | <0.001 |
| glmer(corresponding_author_female ~ year + (1\|group_one), data = cochrane, family = binomial, control = glmerControl(optimizer = "bobyqa", optCtrl=list(maxfun=2e5)), nAGQ = 10)  AIC: 27695  Random effect variance (SD): 0.187 (0.4321) | | | | |
| year | 0.023 | 1.023 | 0.002 | <0.001 |
| Intercept | -46.501 | <0.001 | 4.777 | <0.001 |
| glmer(last_author_female ~ year + (1\|group_one), data = cochrane, family = binomial, control = glmerControl(optimizer = "bobyqa", optCtrl=list(maxfun=2e5)), nAGQ = 10)  AIC: 26735  Random effect variance (SD): 0.203 (0.4501) | | | | |
| year | 0.017 | 1.017 | 0.002 | <0.001 |
| Intercept | -33.741 | <0.001 | 4.997 | <0.001 |
